# Genetic variants disrupting activity-dependent CELF2 shuttling cause neuronal hyperexcitability, learning deficits, and seizures

**DOI:** 10.1101/2025.06.20.25329512

**Authors:** Michelle Hua, Mohamad-Reza Aghanoori, Melissa MacPherson, Yi Ren, Yifan Yang, Yan Yan Or, Laura Williams, Christopher J. Gafuik, Chloe Quelin, Boris Keren, Sarah Schuhmann, Georgia Vasileiou, Alexia Bourgois, Antonio Vitobello, Christophe Philippe, Zornitza Stark, Richard J. Leventer, Frederic Tran-Mau-Them, Marine Tessarech, Clément Prouteau, Phillis Lakeman, Mahdi Motazacker, Donald R. Latner, Raymond Caylor, Eloise Prijoles, Angie Lichty, Yvette van Ierland, David Sweetser, Edward Steel, Jan Cobben, Majed J. Dasouki, Daniel Calame, Gengyi Wang, Brooke Rackel, James Ellis, Guiqiong He, Douglas J. Mahoney, Micheil Innes, Jonathan Epp, Guang Yang

## Abstract

*De novo* heterozygous variants in the *CELF2* gene have recently been associated with a rare neurodevelopmental disorder. However, the mechanisms linking specific variants to distinct clinical phenotypes remain poorly understood. Here, we report a new cohort of 14 individuals with *de novo CELF2* variants, providing evidence that variants causing CELF2 cytoplasmic mislocalization, but not its loss-of-function, are associated with seizures. Using proband induced pluripotent stem cell-derived neurons and transgenic mouse models, we show that CELF2 undergoes activity-dependent nucleocytoplasmic shuttling in excitatory neurons, and its cytoplasmic retention causes neuronal hyperexcitability, leading to learning and memory deficits. In the cytoplasm, CELF2 regulates mRNAs critical for synaptic functions and neuronal excitability implicated in epileptic seizures and intellectual disability. Through drug screening, we identify AKT signaling as a key regulator of CELF2 shuttling and a target for treating CELF2-associated hyperexcitability. Our findings expand the clinical and genetic spectrum of CELF2-related neurodevelopmental disorders and reveal variant-specific mechanisms that link CELF2 mislocalization to neuronal hyperexcitability, learning deficits, and epileptic seizures.

**One Sentence Summary:** *CELF2* variants link protein mislocalization to neuronal hyperexcitability, learning deficits, and epileptic seizures.

## INTRODUCTION

The mammalian CUGBP Elav-like family member 2 (*CELF2*) gene encodes a highly conserved multifunctional RNA-binding protein (RBP)(*1*). CELF2 contains three RNA recognition motifs (RRMs) and multiple regions resembling nuclear localization (NLSs) and export signals (NESs), which allow it to shuttle between the nucleus and cytoplasm and engage in multiple facets of RNA regulation (*1–3*). In the nucleus, CELF2 proteins bind to introns in pre-mRNA to control alterative splicing, whereas in the cytoplasm they promote or inhibit mRNA stability and translation in a target-specific manner (*4–7*). Therefore, alterations in CELF2’s subcellular distribution (*7*), expression (*8*), and RNA-binding activity (*9*) can potentially have varying effects on gene expression, leading to distinct functional outcomes. In this regard, we and others have previously reported individuals with *de novo* heterozygous missense variants that are exclusively clustered within and disrupt C-terminal NLSs, causing CELF2 mislocalization from the nucleus to cytoplasm without affecting its expression (*7, 10*). These individuals all have a neurodevelopmental disorder (NDD) with features that may include seizures, global developmental delay, intellectual disability (ID), speech and language impairment, and autism spectrum disorder (ASD), and brain malformations. Although the C-terminal NLSs partially overlap with RRM3 that is involved in RNA binding (*3*), mislocalized CELF2 variants do not appear to disrupt their RNA-binding or splicing ability (*7*). This suggests that the observed patient phenotypes are unlikely to result solely from a loss-of-function (LoF) mechanism. Instead, they may reflect a combination of cytoplasmic gain-of-function (GoF), nuclear LoF, and/or other combinatorial effects due to CELF2 mislocalization, ultimately leading to phenotypic diversity. However, clinical and experimental evidence supporting the specific mechanisms of action of these variants in driving distinct phenotypic features is still lacking. *CELF2* is expressed throughout brain development, particularly in forebrain structures such as the cerebral cortex (*7, 11, 12*). Beyond its diverse effects on RNA, CELF2’s pleiotropic roles in brain development and the cell-type specific impacts of the variants also likely contribute to the broad spectrum of neurodevelopmental symptoms. Our previous work demonstrates that in the embryonic mouse cortex, the dynamic nucleocytoplasmic shuttling of CELF2 is critical for maintaining neural precursor cell (NPC) homeostasis (*7*). By coordinating the translation of pro-differentiation mRNAs, CELF2 balances NPC proliferation and neurogenic differentiation, and mislocalization of mutant CELF2 disrupts this balance, causing abnormal development of the cortex (*7*). While these NPC defects may help explain certain features of cortical malformation such as macrocephaly and abnormal gyral patterns, other clinical phenotypes such as seizures, ID, and related learning deficits point to additional pathogenic mechanisms likely involving dysregulation of mature neurons (*13, 14*). In this regard, upon differentiation from NPCs, wild-type (WT) CELF2 relocates predominantly to the nucleus in newborn neurons, with its expression increasing substantially in excitatory neurons in the postnatal brain (*7, 15*). Further supporting the notion, a key feature of epilepsy is the hyperexcitability of excitatory neurons and their synchronized firing (*14*), often driven by pathological changes in genes encoding ion channels (e.g., KCNA1/2)(*16*), glutamate transporters (e.g., SLC1A2)(*17*), and synaptic proteins (e.g., STX1B)(*18*). Neuronal hyperexcitability can also disrupt synaptic dynamics and coordinated activity of neuronal ensembles critical for encoding memory traces, leading to learning deficits involved in ID (*19–21*). Intriguingly, *CELF2* knockout (KO) mice show normal spatial learning and memory but exhibit ASD-like behaviors, accompanied by reduced dendritic spine density and impaired synaptic maturation (*8*). These findings further suggest that CELF2 mislocalization-induced phenotypes may arise from mechanisms beyond LoF alone. However, the limited understanding of how CELF2 functions in neurons and the lack of information to stratify patient phenotypes make it difficult to ascertain the mechanisms that link CELF2 mislocalization to specific clinical presentations.

Here, we describe 14 individuals with *de novo* heterozygous *CELF2* variants, including new and recurring missense variants and protein-truncating variants (PTVs) that differentially cause CELF2 mislocalization and/or LoF. Phenotypic and functional assessments revealed that variants causing CELF2 mislocalization but not LoF are associated with the presentation of seizures. Using genetically engineered mice, we found that CELF2 mislocalization but not LoF caused neuronal hyperexcitability in the cortex and hippocampus, resulting in network hyperactivity, failed separation of neuronal ensembles for learning tasks, and memory deficits in the fear conditioning test. Mislocalized CELF2 proteins bound mRNAs encoding key regulators of synaptic functions and neuronal excitability implicated in epilepsy and intellectual disability. Interestingly, we found that CELF2 translocates between the nucleus and cytoplasm in response to neuronal activity, suggesting its dynamic shuttling regulates activity-dependent plasticity of intrinsic excitability, with mislocalization variants disrupting this process and causing hyperexcitability. Cell-based drug screening revealed AKT signaling in promoting cytoplasmic CELF2 translocation, and AKT inhibition reversed the hyperexcitability defects in patient-derived cortical neurons (iNs). Our findings expand the clinical and genetic spectrum of CELF2-related neurodevelopmental disorders and highlight a critical role of dynamic CELF2 shuttling in regulating neuronal excitability and brain functions. Moreover, our results indicate that variants disrupting CELF2 function in diverse ways can lead to distinct clinical manifestations, underscoring the need for tailored therapeutic strategies.

## RESULTS

### Missense variants causing CELF2 mislocalization are associated with seizures, whereas PTVs are not

We and others previously identified heterozygous variants that perturb CELF2’s C-terminal NLSs (*7, 10*). Using the GeneMatcher platform (*22*), we subsequently identified an additional 14 individuals with heterozygous missense variants and PTVs across the *CELF2* gene, including two previously described recurring missense variants (p.Arg493His, p.Pro507Ser) (**Figure 1A**). The genetic and clinical data are summarized in **Table S1**. All variants were *de novo* and comprised six LoF variants (nonsense/frameshift), one frameshift variant predicting truncated CELF2 (p.Gly414AlafsTer45), and seven missense variants, of which three were located within the C-terminal NLSs, two within the divergent domain between RRM2 and RRM3, and two in the N-terminal region, either within or adjacent to RRM1. One fetal case (p.Asn26Thr) exhibited hydrocephalus and agenesis of the corpus callosum with moderate ventricular dilatation. The other 13 individuals in the cohort ranged in age from early childhood to adulthood with no significant sex differences (54.4% females and 45.6% males). These individuals were described as having distinctive and overlapping NDD features, including developmental delay/intellectual disability, speech delay, and epileptic seizures. ASD and other behavioral disorders were also diagnosed. Although NDD features were often observed within this cohort, we noticed that seizures were present exclusively in individuals with missense variants but were absent in those with PTVs **(Figure 1B**). The inclusion of previously identified variants in the analysis further reinforced the potential connection between CELF2 missense variants with seizure manifestations (**Figure S1A**).

**FIGURE 1.**
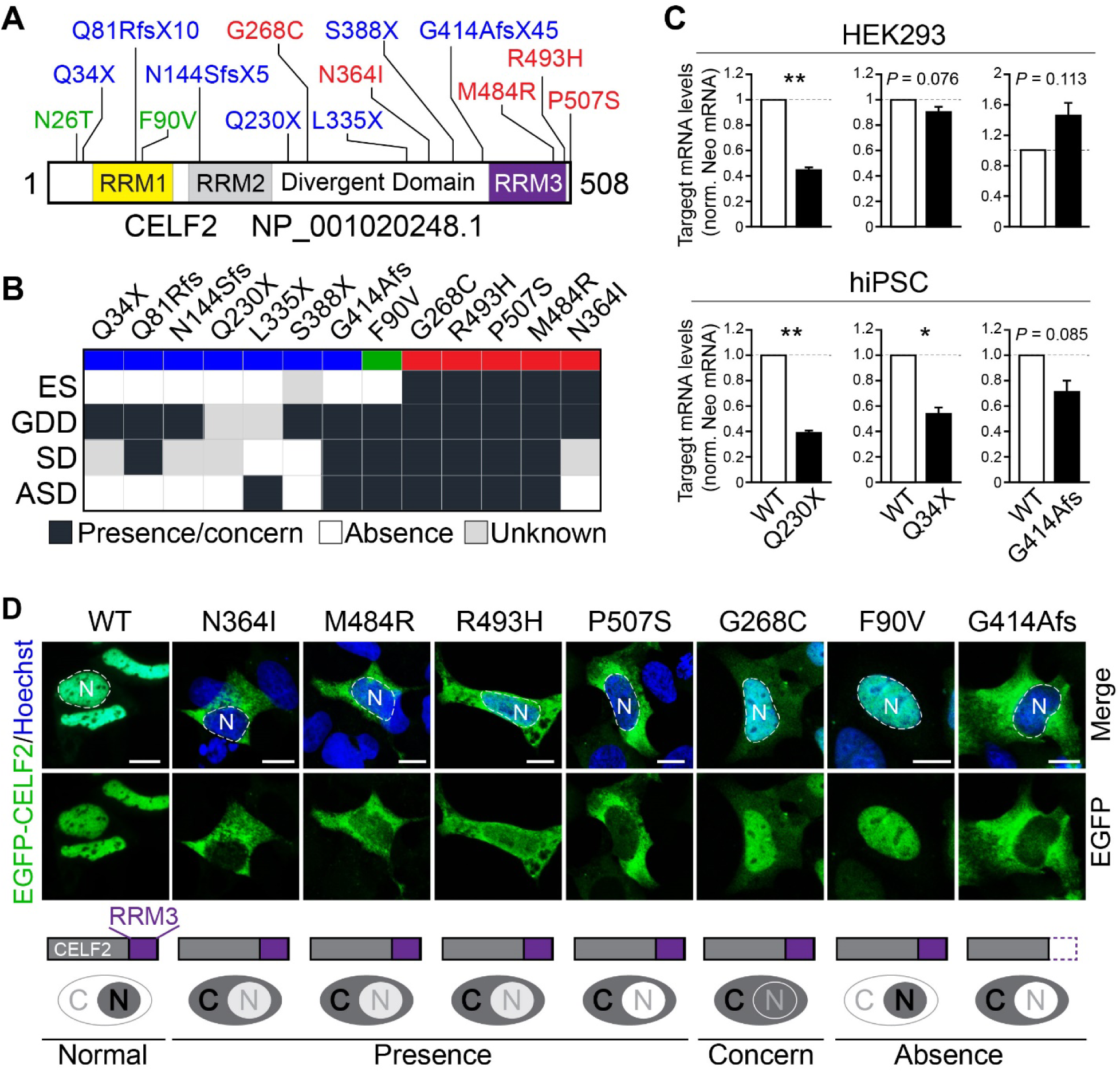
CELF2 missense variants causing CELF2 mislocalization, but not PTVs, are associated with the presence of epileptic seizures. **(A)** Schematic showing the positions of identified variants in the CELF2 protein, containing three RNA-recognition motifs (RRM1-3). PTVs are shown in blue, missense variants causing CELF2 mislocalization in red, and other missense variants in green. **(B)** Heatmap of identified variants and associated clinical features in corresponding individuals, including ES (epileptic seizures), GDD (global developmental delay), SD (speech delay) and ASD (autism spectrum disorder). The presence of a feature is indicated by a black square, and a grey box denotes unavailable information. **(C)** Bar graphs showing normalized mRNA levels of minigene reporters with the indicated variants, compared to WT, in HEK293 cells or human iPSCs, as determined by RT-qPCR. Student’s t-test. *, P < 0.05; **, P <0.01. **(D)** Confocal images of HEK293 cells expressing WT EGFP-CELF2 (green) or that with the indicated variants. Nuclei were counterstained for Hoechst 33258 (blue) and are outlined with dashed white lines. ‘‘N’’ denotes the nucleus. Schematics of the CELF2 protein with or without RRM3 based on the variant’s effect are shown at the bottom, alongside diagrams summarizing the subcellular distribution of EGFP-CELF2 between the nucleus (“N”) and cytoplasm (“C”), as well as the presence or absence of epileptic seizures (EC) in corresponding individuals. Scale bars, 5 μm.

All the identified PTVs are predicted to trigger nonsense-mediated mRNA decay (NMD), except p.Gly414AlafsTer45 which introduces a premature stop codon near the end of the second-to-last exon (**Figure 1A**). To test this, we expressed minigene reporters for several PTVs in human embryonic kidney 293 (HEK293) cells. RT-qPCR analysis showed a significant reduction in p.Gln230Ter reporter mRNA levels compared to WT, confirming its NMD-activating effect, whereas p.Gln34Ter appeared to escape from NMD (**Figure 1C**). However, in human induced pluripotent stem cells (iPSCs), both variants reduced reporter mRNA levels (**Figure 1C**), suggesting cell type-specific NMD activation. As expected, p.Gly414AlafsTer45 did not affect reporter mRNA levels in either cell type (**Figure 1C**), consistent with NMD escape and the production of truncated CELF2 proteins lacking the entire RRM3 and NLSs in the C-terminal region (**Figure 1A**).

To examine the effects of missense variants, we expressed each variant fused to the C-terminus of EGFP in HEK293 cells. While WT CELF2 was predominantly present in the nucleus, all variants except p.Phe90Val and p.Asn26Thr showed varying degrees of cytoplasmic localization, which correlated with the presence/concern of seizures in the individual carrying the mislocalization variants (**Figure 1D, S1B**). In contrast, although the truncated CELF2 p.Gly414AlafsTer45 variant was similarly mislocalized to the cytoplasm, the affected patient did not experience seizures (**Figure 1D**), raising the possibility that a functionally intact RRM3, alongside cytoplasmic mislocalization, may be related to the pathogenesis of epileptic seizures in the patients.

### Cytoplasmic mislocalization of CELF2 causes hyperexcitability of neurons

Seizures are characterized by hyperexcitability of neurons (*14*). To explore the mechanistic links between CELF2 mislocalization and seizures, we leveraged iPSCs derived from a patient carrying a recurring mislocalization missense variant (p.Arg493His) and isogenic iPSCs in which the variant was corrected using CRISPR/Cas9-based gene editing (*23*). Immunostaining showed a substantially higher level of cytoplasmic CELF2 in p.Arg493His iPSCs compared to both isogenic iPSCs and control iPSCs derived from a healthy individual (**Figure 2A, B, S2A, B**). This was corroborated by subcellular fractionation and Western blot (WB) analysis, which showed an 8-fold increase in the cytoplasmic-to-nuclear ratio of CELF2 in p.Arg493His iPSCs (**Figure 2C, D**, **S2C**).

**FIGURE 2.**
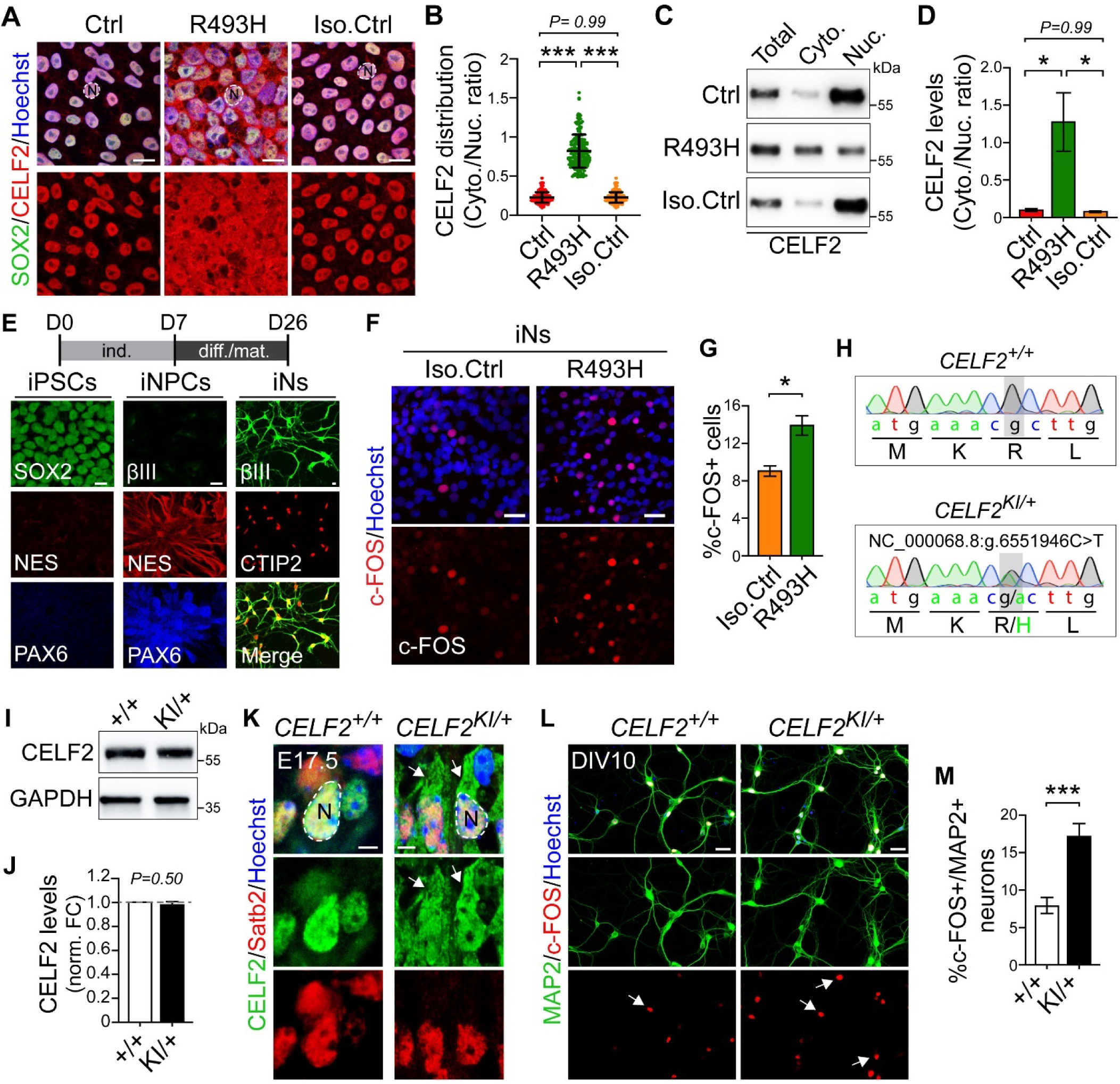
The p.R493H variant causes CELF2 mislocalization and neuronal hyperexcitability. **(A)** Confocal images of control (Ctrl), p.R493H proband, and isogenic control (Iso. Ctrl) iPSCs, immunostained for CELF2 (red) and the iPSC marker Sox2 (green). **(B)** Quantifications of CELF2 distribution in iPSCs as shown in (A). **(C)** Western blots of total lysates, cytoplasmic (Cyto.), and nuclear (Nuc.) fractions from the indicated iPSCs, probed for CELF2. **(D)** Quantifications of the cytoplasmic-to-nuclear ratio of CELF2 levels in iPSCs from (C). **(E)** Schematic and confocal images showing the differentiation of iPSC into iNPCs and iNs at day 7 (D7) and D26 post-induction, immunostained for corresponding markers as indicated. **(F)** Confocal images of p.R493H and isogenic control iNs immunostained for c-FOS (red). **(G)** Quantifications of c-FOS-positive cells in iN cultures from (F). **(H)** Sanger sequencing confirming the heterozygous G>A mutation, resulting in an Arg-to-His substitution (p.R493H) in knock-in (KI) *CELF2^KI/+^* mice. **(I)** Western blots of cortical lysates from E17.5 embryos, probed for CELF2 and GAPDH as a loading control. **(J)** Quantifications of CELF2 protein levels from (I). **(K)** Confocal images of E17.5 cortical sections, immunostained for CELF2 (green) and the excitatory neuronal marker Satb2 (red). Arrows highlight CELF2 signals in the cytoplasm. **(L)** Confocal images of cultured iNs immunostained for c-FOS (red) and the neuronal marker MAP2 (green). Arrows point to c-FOS-positive neurons. **(M)** Quantification of c-FOS-positive cells relative to total neurons in iN cultures as shown in (L). Nuclei were counterstained with Hoechst 33258 (blue in A, F, K, L) and are outlined with dashed white lines in (A, K) with ‘‘N’’ denoting the nucleus. Data are presented as means ± SEM. One-way ANOVA with Turkey’s test (B and D) and Student’s t-test (G, J, and M). *, P < 0.05; ***, P <0.001. Scale bars: 2.5 μm in (K), 10 μm in (A, E), 50 μm in (F, L).

We then differentiated iPSCs using the dual SMAD inhibition protocol and confirmed the expression of key marker genes at each stage, including NESTIN (NES) and PAX6 for forebrain NPCs (iNPCs) and βIII-tubulin (βIII) and CTIP2 for excitatory neurons (iNs) (**Figure 2E**). Immunostaining of mature neurons for c-FOS, a marker of neuronal activation, revealed over 50% more c-FOS-positive cells in p.Arg493His iN cultures compared to the isogenic control (**Figure 2F, G**), indicating increased excitability of p.Arg493His iNs.

To further investigate the effects of CELF2 mislocalization on neuronal excitability, we generated knock-in mice harboring one allele with the same p.Arg493His variant using the CRISPR/Cas9 approach (**Figure 2H, S2D**). WB analysis of cortical tissues from *CELF2^R493H/+^* mice (hereafter referred to as “KI”) showed no difference in CELF2 protein levels (**Figure 2I, J**). However, we observed robust cytoplasmic CELF2 in Satb2-positive cortical excitatory neurons at embryonic day (E) 17.5, in contrast to the predominantly nuclear localization in their WT littermates (**Figure 2K**). This aligns with the mislocalization-inducing effect of the p.Arg493His variant. We then prepared primary neuronal cultures from the E17.5 cortex and performed immunostaining after 14 days *in vitro* (DIV), when neurons become mature enough to fire action potentials and show synchronized network activity (*24, 25*). In KI cultures, we detected twice as many c-FOS-positive neurons as WT cultures (**Figure 2L, M**), recapitulating the findings in p.Arg493His iNs and indicating increased neuronal excitability.

### CELF2 mislocalization but not LoF causes neuronal hyperexcitability *in vivo*

To ask whether CELF2 mislocalization affects excitatory neurons in their native environment, we performed *in vivo* analyses of KI mouse brains. Consistent with previous reports (*7, 15*), we found that in the postnatal cortex and hippocampus, CELF2 was highly expressed in excitatory neurons immune-positive for Satb2 or Ctip2 (**Figure 3A, B**), but low in inhibitory neurons labeled by γ-aminobutyric acid (GABA) or by TdTomato in a Vgat-Cre reporter line (**Figure 3C, S3A**), which aligns with the cell-autonomous effect of CELF2 mislocalization observed in cultured neurons (**Figure 2F, L)**. Immunostaining analysis at postnatal day (P) 30 revealed a 2∼3-fold increase in the numbers of c-FOS-positive neurons in the KI cortex and hippocampus compared to WT littermates (**Figure 3D-G**). This increase was not due to a change in cell numbers, as WT and KI mice showed no significant difference in cortical thickness and cell density (**Figure S3B-D**). Furthermore, at P45, the young adult stage when inhibitory neuronal circuits reach greater maturity (*26–28*), we likewise observed a robust increase in c-FOS-positive neurons in both male and female KI mice (**Figure S3E-J**), suggesting alterations in the intrinsic firing properties of excitatory neurons.

**FIGURE 3.**
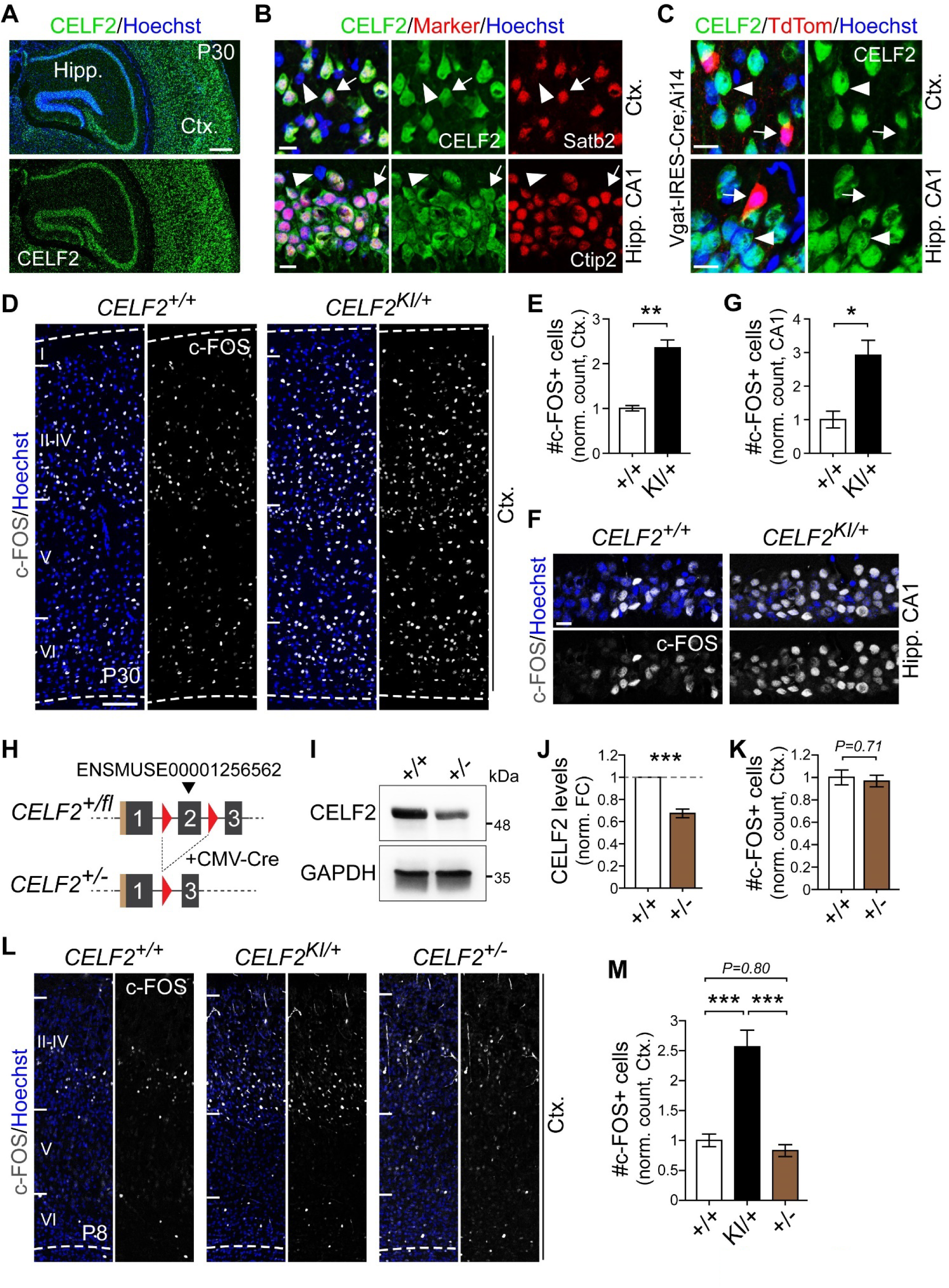
CELF2 mislocalization but not LoF causes neuronal hyperexcitability *in vivo*. **(A)** Confocal images of cortical (Ctx.) and hippocampal (Hipp.) regions from P30 coronal brain sections, immunostained for CELF2 (green). **(B)** Magnified views from (A) showing CELF2 (green) in excitatory neurons, marked by Satb2 or Ctip2 (red). Arrows denote the presence of the markers and arrowheads denote absence. **(C)** Confocal images of brain sections from a Vgat-IRES-Cre; Ai14 reporter mouse, showing TdTomato-labeled inhibitory neurons (red, arrows) and the absence of CELF2 (green) in these cells. **(D-G)** Confocal images (D, F) of P30 cortical sections immunostained for c-FOS (white), with quantifications (E, G) of c-FOS-positive cells in the cortical and hippocampal CA1 regions of *CELF2^KI/+^* mice, normalized to WT *CELF2^+/+^* mice. **(H)** Schematic showing the deletion of exon 2 in the CELF2 gene, flanked by loxP sites (red triangles), by crossing a CMV-Cre deleter with a conditional knockout line (*CELF2^+/fl^*). **(I)** Western blots of cortical lysates from E17.5 WT and heterozygous knockout (*CELF2^+/-^*) mice, probed for CELF2 and GAPDH as a loading control. **(J)** Quantifications of CELF2 protein levels from (I). **(K)** Quantifications of c-FOS-positive cells in the P45 cortex of *CELF2^+/-^* mice, normalized to WT *CELF2^+/+^*mice. **(L, M)** Confocal images (L) of P8 cortical sections immunostained for c-FOS (white), with quantifications (M) of c-FOS-positive cells. Nuclei were counterstained with Hoechst 33258 (blue in A-D, F, L). Data are presented as means ± SEM. Student’s t-test (E, G, J, K) and One-way ANOVA with Turkey’s test (M). *, P < 0.05; **, P < 0.01; ***, P <0.001. Scale bars: 10 μm in (B, C, F), 100 μm in (D, L), 500 μm in (A).

Neuronal hyperexcitability may be driven by nuclear CELF2 LoF, cytoplasmic GoF, or a combination of both, as a result of CELF2 mislocalization. To explore these possibilities, we generated *CELF2* heterozygous knockout mice (*CELF2^+/-^*), which exhibited a roughly 40% reduction in CELF2 protein levels (**Figure 3H-J**). Intriguingly, the number of c-FOS-positive neurons in the *CELF2^+/-^* cortex remained unchanged at P45, compared to WT (**Figure 3K, S3K**). Cortical excitatory neurons undergo rapid growth during early postnatal weeks, with c-FOS first detectable around P8 and steadily increasing over time (**Figure S3L, M**). Notably, even at P8, KI mice already showed a higher number of c-FOS-positive neurons in the cortex, whereas no differences were seen between *CELF2^+/-^*and WT mice (**Figure 3L, M**). These findings suggest that CELF2 mislocalization-induced changes in neuronal excitability are unlikely to be due to a reduction in nuclear CELF2 levels but rather result from its cytoplasmic GoF. This is consistent with the potential involvement of functionally intact CELF2 in the manifestation of seizures in patients with mislocalization variants (**Figure 1D, S1A**).

### CELF2 shows dynamic nucleocytoplasmic distribution during excitatory neuron maturation

To investigate the role of cytoplasmic CELF2 in neuronal excitability, we examined its subcellular distribution in excitatory neurons. In the embryonic mouse cortex, CELF2 was predominantly nuclear (**Figure 2K**). Surprisingly, in the P45 cortex and hippocampus, its localization varied, with some neurons retaining nuclear CELF2 while others showed varying degrees of cytoplasmic localization (**Figure 4A**). A similar pattern was observed in the human brain, where CELF2 also exhibited differential cytoplasmic localization in adult cortical and hippocampal neurons (**Figure S4A, B**), distinct from the embryonic stage (**Figure S4C**). These findings indicate that CELF2 undergoes a localization shift during neuronal maturation. Indeed, analysis of cortical neurons from mid-embryonic to postnatal stages revealed an apparent transition around P5, with CELF2 shifting from predominantly nuclear to cytoplasmic, peaking at around P8 (**Figure 4B, C**). This was then followed by a gradual divergence, with some neurons maintaining cytoplasmic CELF2 while others exhibiting more nuclear localization (**Figure 4B, C**). The initial transition in CELF2 localization during the perinatal stage was further corroborated by cytoplasmic and nuclear fractionation and WB analysis, which showed a marked increase in cytoplasmic CELF2 levels (**Figure 4D-F**). Interestingly, neurons in different cortical layers exhibited varied change in CELF2 distribution (**Figure S4D**), potentially reflecting differences in their maturation rates.

**FIGURE 4.**
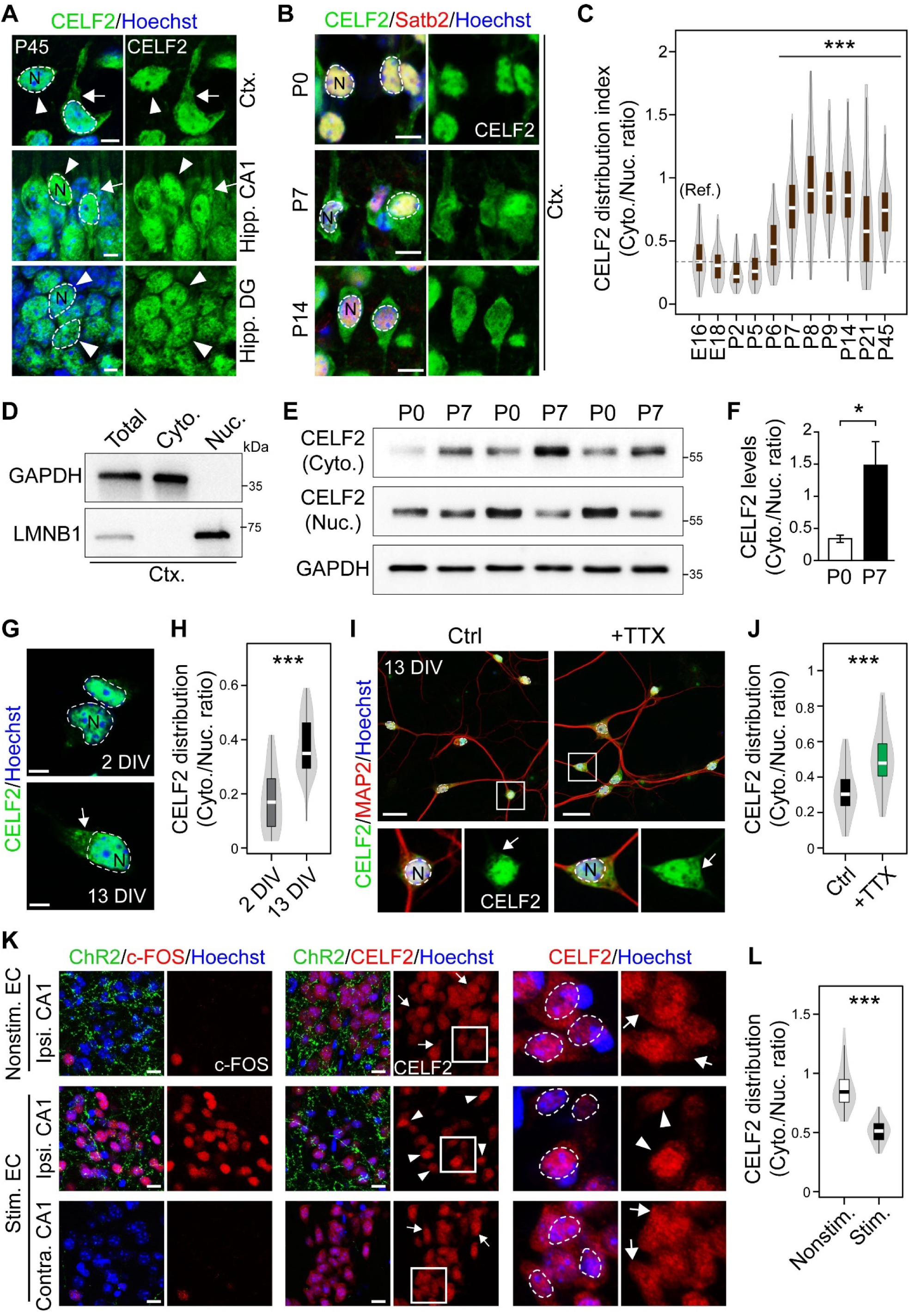
CELF2 shows dynamic subcellular localization in response to neuronal activity. **(A)** Confocal images of cortical and hippocampal regions from P45 mouse brain sections, immunostained for CELF2 (green). **(B)** Confocal images showing CELF2 expression (green) in cortical excitatory neurons, marked by Satb2 (red) from P0, P7, and P14 mouse brains. **(C)** Quantifications of the cytoplasmic-to-nuclear ratio of CELF2 levels in cortical neurons at indicated developmental stages. **(D)** Western blots of total lysates (total), cytoplasmic (Cyto.), and nuclear (Nuc.) fractions from P7 cortical tissues, probed for GAPDH (cytoplasmic marker) and LMNB1 (nuclear marker). **(E)** Western blots of cytoplasmic and nuclear fractions from P0 and P7 cortical tissues, probed for CELF2 and GAPDH. **(F)** Quantifications of CELF2 levels from (E). **(G)** Confocal images of primary cortical neurons cultured for 2 days in vitro (DIV) or 13 DIV, immunostained for CELF2 (green). **(H)** Quantifications of CELF2 distribution in neurons as shown in (G). **(I)** Confocal images of 13 DIV neurons treated with or without TTX for 48 hours, immunostained for CELF2 (green) and MAP2 (red). Magnified views of the white boxed area are shown below. **(J)** Quantification of CELF2 distribution in neurons as shown in (I). **(K)** Confocal images of hippocampal CA1 regions ipsilateral or contralateral to AAV injection sites, with or without blue light stimulation. Sections were immunostained for EGFP-ChR2 (green) and c-FOS or CELF2 (both red). Magnified boxed areas are shown to the right. **(L)** Quantification of CELF2 distribution in CA1 cells as shown in (K). Nuclei were counterstained with Hoechst 33258 (blue in A, B, G, I, K) and are outlined with dashed white lines, with ‘‘N’’ denoting the nucleus. Arrows in (A, G, I, K) highlight cytoplasmic CELF2 while arrowheads point to neurons with higher nuclear localization. Data are presented as means ± SEM. Mann-Whitney U test (C) and Student’s t-test (F, H, J, L). *, P < 0.05; ***, P < 0.001. Scale bars: 5 μm in (A, G), 10 μm in (B, K), 25 μm in (I).

### CELF2 changes subcellular localization in response to neuronal activity

Of note, the initial nuclear-to-cytoplasmic shift of CELF2 localization in cortical neurons preceded the onset of c-FOS expression, and its peak and subsequent divergence coincided with the increasing number of c-FOS-positive neurons (**Figure 4C, S3L, M**). Moreover, mature neurons in 14 DIV cultures showed higher cytoplasmic CELF2 levels compared to immature neurons at 2 DIV (**Figure 4G, H**). These findings raise the possibility that CELF2 may change its localization in response to neuronal activity to modulate intrinsic excitability. To test this, we treated 14 DIV primary neuronal cultures isolated from the mouse cortex with tetrodotoxin (TTX), a sodium channel blocker, to inhibit neuronal activity for 48 hours. This treatment led to a significant increase in CELF2 cytoplasmic localization (**Figure 4I, J**). Next, to ask whether neuronal activation induces the opposing effect to drive CELF2 translocation into the nucleus, we used an optogenetics approach to photo-stimulate entorhinal cortex (EC) afferents, which project (both directly and indirectly) to and activate CA1 pyramidal neurons in the hippocampus. To this end, adult mice were injected with an AAV viral construct encoding the light-activated ion channel channelrhodopsin-2 (ChR2-EGFP) into the lateral EC, followed 2 weeks later by pulsed blue light stimulation over a period of 5 minutes. Immunostaining analysis of tissues 90 minutes post-stimulation confirmed proper ChR2-EGFP expression in EC fibers within ipsilateral CA1 regions, alongside robust c-FOS induction in pyramidal neurons upon photo stimulation. Importantly, these hyperactivated neurons showed greater nuclear CELF2 localization compared to non-stimulated neurons or those in the contralateral CA1 lacking ChR2-EGFP fibers (**Figure 4K, L**). Our results show that cytoplasmic CELF2 promotes neuronal excitability and its nuclear translocation in activated neurons may enable their adjustment of excitability. Disrupting this dynamic shuttling of CELF2 leads to hyperexcitability.

### Cytoplasmic CELF2 binds mRNAs implicated in epileptic seizures and regulators of neuronal excitability

To understand how cytoplasmic CELF2 accumulation induces hyperexcitability relevant to seizures, we sought to identify CELF2 target mRNAs in the cytoplasm. We performed RNA immunoprecipitation-sequencing (RIP-seq) with a specific antibody against CELF2 for cytoplasmic RNA isolated from the P0 KI cortex, a timepoint when CELF2 is normally in the nucleus of WT neurons (**Figure S5A, 4B, C**). Three biological replicates of CELF2 RIPs identified 1,192 protein-coding mRNAs as targets enriched by at least two-fold compared to control IgG RIPs and total input samples, with a false-discovery rate (FDR) < 0.01 (**Figure 5A, S5B, Table S2**). Human Phenotype Ontology analysis of these target mRNAs revealed a specific enrichment of terms related to epilepsy/seizures and ID (**Figure 5B**), consistent with observed patient phenotypes. Examples of genes associated with these terms included voltage-gated ion channels (e.g., *Kcna2*, *Scn3a*), ion and glutamate transporters (e.g., *Slc12a5*, *Slc1a2*), and regulators of synaptic transmissions (e.g., *Grin2b*, *Syt2*) (**Figure 5C**). Pathogenic variants in these genes are not only associated with epilepsy, seizures, ID, but also neurodevelopmental disabilities including those seen in CELF2-related patients. For instance, GoF mutations in *SCN3A* are associated with epileptic encephalopathy, ID, and cortical malformation (*29, 30*), while *SLC12A5* variants are associated with epilepsy of infancy with migrating focal seizures and profound ID and ASD (*31–33*). Importantly, alterations in the function of these genes have been shown to change neuronal excitability in mice and human iNs (*34–36*), phenocopying CELF2 KI mice and p.R493H patient iNs.

**FIGURE 5.**
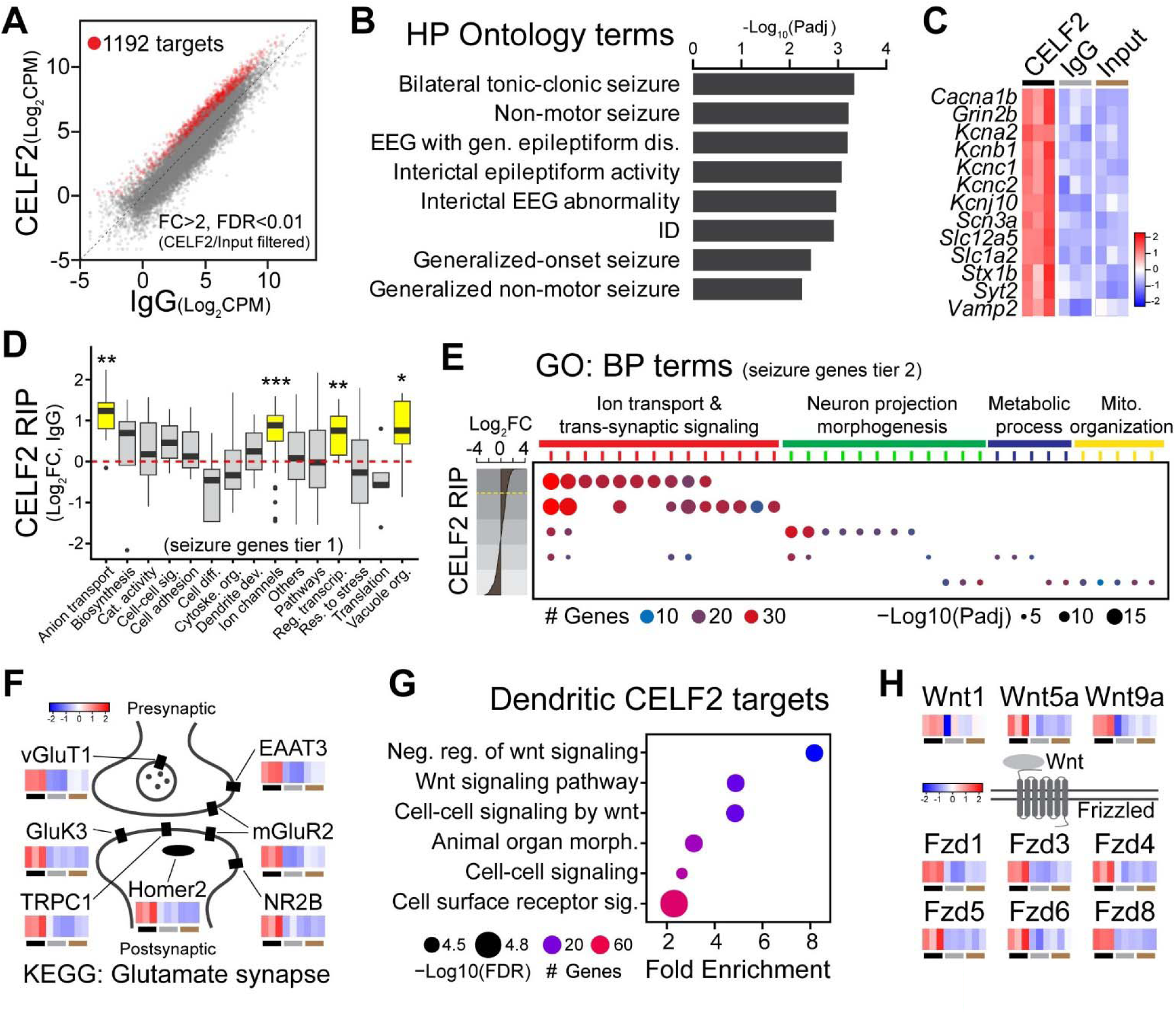
Mislocalized CELF2 in the cytoplasm binds mRNAs of seizure-related genes and regulators of intrinsic excitability. **(A)** Scatterplot showing log_2_-transformed averaged RIP-seq counts per million (CPM) for CELF2 versus IgG. 1,192 genes enriched by at least a 2- fold change with FDR < 0.01 relative to IgG RIP and total input are highlighted in red. **(B)** Human Phenotype (HP) Ontology analysis of 1,192 CELF2 targets, showing the top eight enriched terms with log_10_-transformed adjusted P-values. **(C)** Heatmap showing z-score transformed expression levels of selected genes from the enriched HP ontology terms across CELF2 RIP, IgG RIP and total input samples. **(D)** CELF2 RIP enrichment (log_2_ fold-change of RIP/IgG) for 142 seizure-related genes (Tier 1, Macnee et al., 2023), across 15 functional groups based on Gene Ontology (GO) terms. Groups that show significantly higher enrichment are highlighted in yellow. Mann-Whitney U test. *, P < 0.05; **, P < 0.01; ***, P <0.001. **(E)** Dot plot showing enriched GO terms from the analysis of 514 Tier 2 seizure-related genes, grouped into five bins of equal size based on their CELF2 RIP enrichment, as shown on the left. The yellow dotted line marks a 2-fold change in RIP/IgG. The enriched GO terms are grouped into four Biological Process (BP) categories. Color indicates the number of genes mapped to each term, while dot size represents adjusted p-value. **(F)** Schematic diagram of glutamate synapse-related genes identified through KEGG analysis of CELF2 targets, accompanied by heatmaps showing their z-score transformed expression levels across CELF2 RIP, IgG RIP and total input samples as illustrated in (C). **(G)** Dot plot showing top six enriched GO BP terms, with fold enrichment values, from the analysis of 185 CELF2 target mRNA identified as dendritic mRNA (Hacisuleyman et al., 2024). **(H)** Schematic of Wnt ligands and Frizzled receptors identified as CELF2 targets, with heatmaps showing their z-score expression levels as shown in (C).

To ask which molecular processes may mediate the collective contribution of CELF2 target mRNAs to neuronal excitability and epilepsy, we examined known epilepsy-associated genes, which cause seizures through different mechanisms (*37*). We found only a slight, yet significant, enrichment of 143 high-confidence epilepsy genes (tier 1, Macnee 2023) in CELF2 RIPs, compared to randomly selected control genes (**Figure S5C**). This suggests that CELF2 may influence some, but not all, seizure-related pathways. Indeed, functional grouping revealed that the enriched genes were specifically associated with ion transport, ion channels, transcription, and vacuole organization, but not with pathways related to cell differentiation, catalytic activity and others (**Figure 5D**). Further analysis of 738 broader epilepsy genes (tier 2, Macnee 2023) also showed that the top-enriched genes were primarily involved in ion transport and synaptic regulation (**Figure 5E**), consistent with their encoded proteins localizing to synapses, dendrites, and membrane structures (**Figure S5D**). In contrast, other tier 2 genes related to neuron structure and metabolism showed no enrichment in CELF2 RIPs (**Figure 5E**). Gene Ontology (GO) and KEGG analysis of CELF2 targets highlighted cell-cell signaling and glutamatergic synapse pathways (**Figure 5F, S5E**), further supporting a direct link of CELF2 to excitatory synaptic signaling. Interestingly, CELF2 has been shown to bind mRNAs regulating neuronal excitability in human cortical organoids (*9*). Despite model differences, we found approximately 10% overlap in CELF2 target mRNAs, linked to the dendritic compartment and glutamatergic synapse (**Figure S5F, G**).

This compartmentalization feature of CELF2 target mRNAs suggests that activity-dependent changes in CELF2’s cytoplasmic availability may influence local mRNA regulation in dendrites to modulate synaptic functions and neuronal excitability. To explore this possibility, we examined dendritic mRNAs identified in hippocampal CA1 neurons (*38*). Approximately 15% of CELF2 target mRNAs were found in dendrites, consistent with GO analysis showing their enrichment in synapse, cell-cell junctions, and somatodendritic compartments, with roles in cell-cell and receptor signaling (**Figure 5G, S5H**). Notably, these included regulators of Wnt signaling pathways, such as Wnt ligands and receptors (**Figure 5H**). Both canonical and non-canonical Wnt signaling are known to regulate intrinsic firing properties, with their dysregulation linked to epilepsy (*39, 40*) and ID (*41–43*). Wnt5a, for instance, can act as an autocrine factor that rapidly modulates synaptic transmission and intrinsic excitability by remodeling spines (*44*) and mobilizing glutamate receptors (*40*). These rapid effects of Wnt5a align with its expression and secretion under the control of local translation in response to synaptic activity (*45–47*). Accordingly, disruption of Wnt5a expression and function has been shown to impair learning and memory in mice (*48*).

### CELF2 mislocalization-induced neuronal hyperexcitability perturbs learning and memory process

Activity-dependent plasticity of intrinsic excitability mediates the formation of neuronal ensembles that underlie learning and memory (*19*). Given the ID and speech delay in patients and the CELF2 mislocalization-induced hyperexcitability, we asked whether disruptions in activity-dependent CELF2 translocation may alter activation patterns of neuronal ensembles critical for learning and memory. To this end, we expressed the genetically encoded calcium indicator GCaMP8f in excitatory pyramidal neurons by injecting AAV9 into the hippocampal CA1 region of WT and KI mice. Using Ca^2+^-activity as a proxy, population neuronal activity was recorded using fiber photometry during contextual fear conditioning protocol (**Figure 6A**). Both the male and female KI mice demonstrated an impairment in retention of the contextual fear memory when assessed 24 hours after acquisition (**Figure 6B**). Examining the photometry signal, we found that during the acquisition phase of the contextual fear conditioning task, both male and female KI mice showed significantly larger population activity in response to the foot shocks compared to their WT littermates (**Figure 6C-E**), consistent with hyperexcitability of KI neurons. Interestingly, despite the hyperactivity during task acquisition, the CA1 population response was significantly reduced during the retrieval test (**Figure 6F, G**), potentially as a correlate of the impaired retrieval. During bouts of movement in the context memory retrieval session we observed decreases in the peak frequency and area under the curve (AUC) of the photometry signal in both male and female KI mice (**Figure 6H, I**). No significant changes were observed in these metrics during bouts of freezing (**Figure S6A-D**).

**FIGURE 6.**
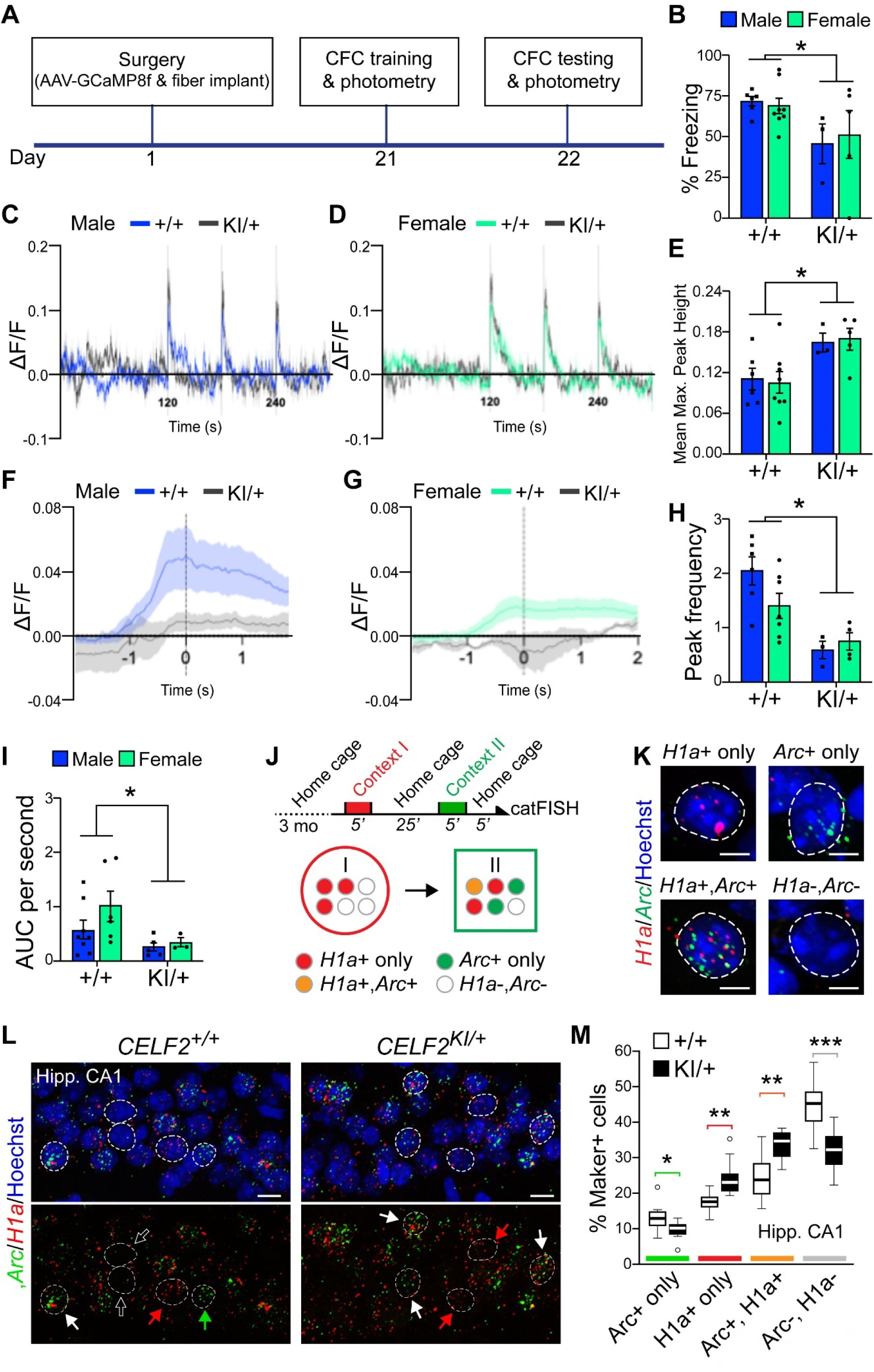
Neuronal hyperexcitability caused by CELF2 mislocalization disrupt learning and memory. **(A)** Schematic diagram illustrating the experimental setup for fibre photometry and contextual fear conditioning (CFC). Three weeks after AAV-mediated GCaMP8f expression and fibre implantation in the CA1 region, neuronal responses were recorded during foot shock trails, followed by retrieval tests. **(B)** KI mice displayed impaired retention of the contextual fear memory compared to WT mice. **(C, D)** Photometry ΔF/F traces recorded during contextual fear conditioning acquisition. Red vertical lines indicate timing of foot shock delivery. **(E)** Quantifications of photometry amplitude in male and female mice, in response to the foot shocks showed increased neuronal activity in KI mice. **(F, G)** Photometry traces during memory retrieval specifically during transitions from freezing to moving (± 2 seconds from the offset of freezing). During these standardized bouts of activity, KI mice showed significantly reduced activity compared to WT mice in both males and females with respect to **(H)** Peak frequency of the photometry signal and **(I)** AUC of the photometry trace. **(J)** Schematic diagram showing the experimental setup for sequential context exploration and catFISH, with four nuclear expression profiles of *Arc* and *H1a* mRNA in response to contexts I and II. **(K)** Confocal images showing representative ISH signals for *H1a* (red) and/or *Arc* (green) mRNA in hippocampal CA1 neurons. **(L)** Confocal images of hippocampal CA1 regions subject to catFISH following context exploration. Red, green, white, and empty arrows indicate H1a-only, Arc-only, double-positive, and IEG-negative neurons, respectively. **(M)** Quantifications of IEG expression in CA1 neurons as shown in (K). Nuclei were counterstained with Hoechst 33258 (blue in G, H) and are outlined with dashed white lines. Data are presented as means ± SEM. Two-way ANOVA with Tukey post-hoc test and Student’s t-test. *, P < 0.05; ***, P < 0.001. Scale bars: 5 μm in (J), 10 μm in (K).

During learning, neurons with higher intrinsic excitability are more likely to be activated and recruited for memory encoding, while a subsequent reduction in excitability of previously activated neuron ensembles ensures the separation of different memories (*19*). Persistent hyperexcitability perturbing these fluctuations has been shown to cause memory impairment (*49*). Indeed, we found that KI mice showed significant reduction in freezing responses during the subsequent memory retrieval after fear-conditioning trials (**Figure 6B**).

To ask if this reflects the aberrant recruitment and separation of neuronal population in the first training session, we examined neuronal population responses to distinct environmental exploration experiences by sequentially exposing naïve mice to contexts A and B for 5 min each, with a 25-min interval (**Figure 6J, S6E**). This paradigm activates distinct subsets of CA1 neurons in each context, inducing immediate-early gene (IEG) transcription at varying rates and subsequent mRNA translocation out of the nucleus. For example, the IEG *Arc* can be activated in less than 5 min and quickly translocated, while *Homer1a* (*H1a*) activation takes 30 min. These distinct temporal dynamics enable precise labeling of neuronal ensembles specific to context A or B, using cellular compartment analysis of temporal activity by fluorescent *in situ* hybridization (catFISH) (*50*). Indeed, in the nucleus we found CA1 neurons with only *H1a* mRNA (earlier activation by context A), *Arc* mRNA (recent activation by context B), both mRNAs (activation by both contexts), and no IEG mRNA (no activation) (**Figure 6K**).

Consistent with elevated neuronal excitability, KI mice showed a significantly greater number of activated CA1 neurons expressing each IEG mRNA compared to WT mice (**Figure 6L, M**), indicating increased recruitment of neurons in response to context exploration. Notably, while KI mice showed an overall increase in total *Arc* mRNA-positive neurons responding to context B, the number of neurons with only *Arc* mRNA was significantly reduced, with a corresponding increase in neurons expressing both *Arc* and *H1a*. This suggests aberrant reactivation of previously activated neuronal populations from context A in response to context B in KI mice (**Figure S6E**). Notably we observed this aberrant increase in overlapping neuronal activity in distinct contexts in area CA1 of the hippocampus but not in the CA3 region or the anterior cingulate cortex (ACC) (**Figure S6F, G**). CA1 has been demonstrated to be essential for context memory formation and retrieval (*51*) and therefore is likely to be highly engaged in the context discrimination task used here. It is likely that other tasks which differentially engage different brain regions would also display enhanced excitability in KI mice.

### Drug screening reveals AKT signaling as a regulator of CELF2 translocation and a potential target for restoring neuronal hyperexcitability

CELF2 contains multiple NLSs and NESs that collectively determine its localization (*3*). To ask whether the mislocalization of mutant CELF2 is reversable, we generated a stable HEK293 cell line expressing the p.Arg493His variant fused to EGFP and carried out high-throughput screening of ∼1,400 small molecule compounds (1 µM, 24 hours) from an FDA-approved Drug Library (Selleckchem) using imaging-based analysis. Immunostaining and subcellular fractionation confirmed cytoplasmic accumulation of p.Arg493His CELF2, in contrast to the predominantly nuclear localization of WT CELF2 in stable cell lines (**Figure S7A-C**). After two rounds of the screening, we identified 8 compounds that significantly restored nuclear levels of p.Arg493His CELF2 (**Figure 7A, S7D**). Triciribine and Omipalisib, which both target the PI3K-AKT pathway, were among the most effective hits. Notably, Triciribine, a selective AKT inhibitor, fully restored mutant CELF2 nuclear localization to WT levels (**Figure 7B**). Treatment of p.Arg493His patient-derived iPSCs with Triciribine similarly lead to a robust nuclear restoration of mutant CELF2 proteins, comparable to isogenic control iPSCs (**Figure 7C, D**). To determine whether AKT signaling drives CELF2 cytoplasmic localization, we treated isogenic control and p.Arg493His iPSCs with the pan-AKT activator SC79, which induced significant cytoplasmic translocation of WT CELF2 and further enhanced the cytoplasmic accumulation of mutant CELF2 (**Figure 7E, F**). These results suggest that CELF2 nucleocytoplasmic shuttling is regulated by AKT-mediated signaling.

**FIGURE 7.**
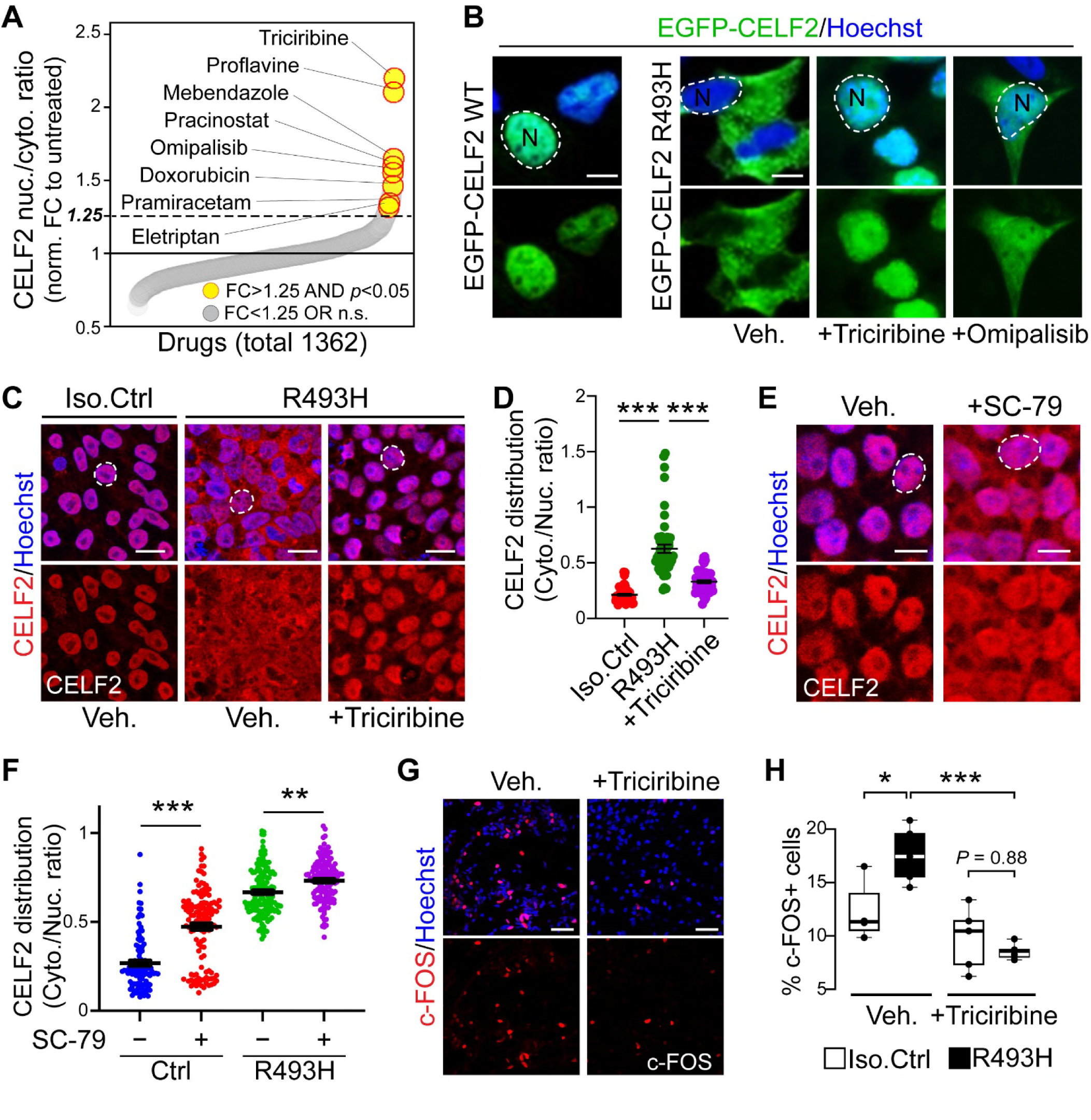
High-throughput drug screening identifies AKT signaling as a regulator of CELF2 translocation and a target for rescuing iN hyperexcitability. **(A)** Drug effect plot showing compounds ranked by their effectiveness in restoring nuclear localization of p.R493H CELF2 in transfected HEK293 cells, compared to the DMSO vehicle. Top-ranked compounds are highlighted in yellow with red circles. **(B)** Confocal images of HEK293 cells expressing EGFP-CELF2 WT or p.R493H mutant (both green), treated with vehicle, Triciribine or Omipalisib. **(C, D)** Confocal images of isogenic control or p.R493H iPSCs (C), treated with vehicle or Triciribine, immunostained for CELF2 (red), and quantifications of CELF2 distribution in iPSCs (D). **(E, F)** Confocal images of control iPSCs (E) treated with vehicle or SC-79, immunostained for CELF2 (red), and quantifications of CELF2 distribution in iPSCs (F). **(G, H)** Confocal images of p.R493H iNs (G) treated with vehicle or Triciribine, immunostained for c-FOS (red), and quantifications of c-FOS-positive cells (H) in isogenic control or p.R493H iNs. Nuclei were counterstained with Hoechst 33258 (blue in B, C, E) and are outlined with dashed white lines, with ‘‘N’’ denoting the nucleus. Data are presented as means ± SEM. One-way ANOVA with Tukey post-hoc test (D, H) and Student’s t-test (F). *, P < 0.05; **, P<0.01, ***, P < 0.001. Scale bars: 5 μm in (B), 10 μm in (C, E), 50 μm in (G).

Given the robust nuclear restoration of mutant CELF2 by AKT inhibition, we next asked whether this could also rescue the associated hyperexcitability defects. To test this, we treated iPSC-derived iNs with triciribine (1 µM, 24 hours) and observed a significant reduction in the excitability of p.Arg493His iNs to the same levels as isogenic control iNs (**Figure 7G, H**). This suggests that the CELF2 mislocalization-induced neuronal hyperactivity are primarily a result of impaired electrical properties rather than growth defects, consistent with our findings on CELF2 target mRNAs (**Figure 6**).

## DISCUSSION

Dysregulation of RBPs contribute to many disorders in humans, but their pleiotropic roles, tissue-specific functions, and complex regulation make it difficult to determine how specific genetic variants in RBPs drive disease pathogenesis, especially in rare diseases with high clinical and genetic heterogeneity. In this study, we focus on *CELF2* variants and provide clinical and mechanistic evidence that mislocalization of functional CELF2 underlies neuronal hyperexcitability, seizures and learning deficits. First, clinical characterization of 14 unrelated individuals with new and recurring *de novo* missense and PTV variants suggests a potential association of mislocalization variants but not LoF variants with epileptic seizures. Second, cortical excitatory neurons derived from patient iPSCs with CELF2 mislocalization showed hyperexcitability, which was phenocopied in KI mice carrying the same variant but not in CELF2 KO mice. Third, neuronal hyperexcitability in KI mice was accompanied by network hyperactivity and behavioral deficits in learning and memory tests, in line with neurodevelopmental deficits seen in patients. Fourth, mislocalized CELF2 can bind a plethora of mRNAs encoding neuronal activity regulators such as ion channels and synaptic signaling modulators that, when mutated, are associated with seizures including epileptic encephalopathy. Notably, we found that CELF2 proteins translocated to the nucleus in response to neuronal activity. Lastly, we showed that the dynamic shuttling of CELF2 is under the control of AKT signaling pathway. AKT inhibition restored nuclear localization of mutant CELF2, thereby rescuing the hyperexcitability defects in patient-derived neurons.

All reported variants to date are heterozygous (*7, 10*), suggesting a dominant model (i.e. haploinsufficiency, GoF, dominant-negative). Our findings also suggest that these variants likely have diverse mechanisms of action, altering CELF2 localization, expression, and/or activity in different ways and, consequently, leading to varied clinical phenotypes. For instance, several nonsense and frameshift variants are predicted to induce LoF via NMD, but their actual effects differ across cell types; while both p.Gln230Ter and p.Gln34Ter triggered NMD in human iPSCs, p.Gln34Ter escaped NMD in HEK293 cells (**Figure 1C**). NMD efficiency is known to vary by cell types, developmental stages, and the position of the premature termination codon (*52, 53*). It is possible that mRNA containing p.Gln34Ter and potentially other PTVs may produce truncated CELF2 lacking different functional domains or act as RNA decoy, disrupting normal CELF2 activity and/or target RNA regulation in a cell-type-specific manner.

On the other hand, mislocalization variants may exert effects beyond nuclear LoF, which aligns with the presence of more clinically significant features in patients, such as seizures and brain malformation (e.g., thin corpus callosum, simplified gyral patterns) (**Table S1**)(*7, 10*). A parallel can be drawn with amyotrophic lateral sclerosis, where mislocalization of the disease-causing RBP TDP-43 results in cytoplasmic GoF acting concurrently with nuclear LoF to drive pathogenesis (*54, 55*). Similar dual pathomechanisms are also seen in other RBPs like FUS (*56, 57*) and hnRNPA2 (*58*). Moreover, cytoplasmic FUS accumulation, rather than nuclear depletion, can selectively induce neuronal defects and social behavioral abnormalities (*59*), highlighting compartment-specific impacts. Likewise, our findings indicate the aberrant neuronal excitability, and the related pathological outcomes are likely driven by cytoplasmic GoF pathomechanisms, involving mislocalized CELF2 with functional RRMs. This is supported by the finding that neuronal hyperexcitability was observed only in KI mice with p.Arg493His

NLS-disrupting variant but was absent in CELF2 KO mice (**Figure 3D-M**), aligning with our observations in patients to date (**Figure 1B**). Although p.Arg493His resides within RRM3, our earlier findings suggest that mutant CELF2 retains its RNA-binding ability, driving phenotypic changes like CELF2 with a WT RRM3 (*7*). We also noticed that a frameshift variant (Gly414AlafxTer45) that escapes NMD but produces a truncated CELF2 lacking the entire RRM3 does not cause seizures despite inducing mislocalization (**Figure 1D**). In contrast, missense variants (p.Gly268Cys and p.Asn364Ile) in the divergent domain, which spare RRM3 but can cause CELF2 mislocalization, are associated with seizures, resembling the effects of NLS-disrupting missense variants. However, it should be noted that discovering more variants in the future will be essential to strengthening the clinical association.

Our finding that CELF2’s RNA-binding activity is potentially linked to seizures suggests that disruptions in the translation, localization, and/or stability of its cytoplasmic target mRNAs underlie aberrant neuronal excitability. Supporting this, cytoplasmic CELF2 binds mRNAs of epilepsy-related genes primarily linked to dysregulation of signal transmission and ion transport, rather than metabolic abnormality, emphasizing its direct impact on synaptic functions and intrinsic excitability (**Figure 5C-E**). Disrupting the expression of these genes, such as the glutamate transporter VGLUT1 (*60, 61*) and potassium channels KCNA2 (*62*) and KCNB1 (*63*) induces neuronal hyperexcitability, mirroring the phenotypes observed in CELF2 KI mice. Many mRNAs associated with synaptic function undergo local translation within dendritic compartments to facilitate rapid and localized modulation of neuronal activity (*64*). We found several CELF2 target mRNAs localized in dendrites, including those encoding Wnt ligands and receptors. These mRNAs have been shown to be extensively regulated at the translational level (*46, 65*) and exhibit expression and localization changes in response to neuronal activity (*45, 66–68*). This activity-dependent regulation enables rapid modulation of Wnt signaling to influence synaptic function and neuronal excitability (*40, 67, 69–72*). Consistent with this, their dysregulation has been linked to epilepsy (*39, 73*). Therefore, persistent retention of mutant CELF2 in the cytoplasm likely occludes the dynamic regulation of its target mRNAs, resulting in impaired synaptic function and altered intrinsic excitability.

An interesting finding of this study is that CELF2 undergoes dynamic shuttling in neurons, with neuronal activation driving its translocation from the cytoplasm into the nucleus. The nucleocytoplasmic shuttling appears to be a general mechanism regulating CELF2 function. For example, cytoplasmic CELF2 translocates into the nucleus to drive neurogenic differentiation in embryonic mouse NPCs (*7*) and a similar transition was seen during chicken heart development (*12*). Conversely, γ-irradiation induces CELF2 nuclear-to-cytoplasmic translocation in human cancer cells (*74*). Although activity-dependent shuttling of RBPs and their regulatory roles in neurons remains underexplored, a few earlier studies provide initial insights. For example, it has been shown that neuronal activation promotes the cytoplasmic translocation of nuclear NOVA, an epilepsy-associated RBP, which allows it to change synaptic protein production and consequently neuronal activity (*75–77*). Similarly, neuronal activation triggers rapid trafficking of two other RBPs, ZBP1 and Sam68, from the cell body into dendrites and spines, allowing local translation of specific synaptic mRNAs (*78, 79*). Our findings here support a model in which CELF2 shuttling facilitates activity-dependent modulation of excitability, which is critical for learning and memory. In this regard, fluctuations in intrinsic excitability may guide the formation of memory-encoding neuronal ensembles, with more excitable neurons preferentially activated to store specific memory (*19*). Subsequent homeostatic mechanisms lower their excitability, allowing other unallocated neurons with increased excitability to encode new memories (*80*). Supporting our model, KI mice showed a greater CA1 population response during the acquisition phase of a contextual fear conditioning task (**Figure 6C-E**). Interestingly,

KI mice demonstrated impaired retention of the fear memory 24 hours later and significantly reduced CA1 activity during the memory retrieval session (**Figure 6B, F-I**), consistent with increased recruitment of neurons because of their aberrantly elevated excitability. The disruption of activity-driven CELF2 nuclear translocation can also impair homeostatic excitability reduction in these neurons. As a result, KI mice showed a significantly greater overlap of neurons activated by exploring distinct contexts (**Figure 6L, M**). Conversely, CELF2 KO mice exhibit normal learning and memory but show social deficits (*8*), further indicating the divergent pathomechanisms contributing to phenotypic heterogeneity in these patients. In light of multiple CELF family members, they are likely to act differentially to regulate neuronal functions, as LoF mutations in CELF4 are otherwise associated with seizures (*81–83*), unlike CELF2 LoF. These differences underscore the needs for tailored patient management and variant-specific therapeutic approaches. These observations also carry implications for syndrome nomenclature. At present, OMIM has associated pathogenic variants in *CELF2* with Developmental and Epileptic Encephalopathy 97 (DEE 97, #619561), although it is plausible that at minimum two distinct NDDs may be associated with heterozygous pathogenic variation in *CELF2*. Using the dyadic approach to syndrome delineation (*84*), we could tentatively consider a CELF2 LoF NDD and a CELF2 related DEE.

What mechanisms regulate CELF2 localization and enable its dynamic shuttling in response to neuronal activity? Deletion of C-terminal NLSs causes cytoplasmic localization, but this effect can be reversed by simultaneously removing N-terminal NES motifs (*3*), which suggests that CELF2 localization is determined by the combinatorial actions of opposing nuclear import and export processes, potentially under the control of specific cellular signaling mechanisms.

Supporting this notion, our small molecule screening and other evidence implicate AKT signaling in this regulation. First, PI3K/AKT inhibition induces nuclear translocation of mutant CELF2, whereas AKT activation promotes CELF2 cytoplasmic localization (**Figure 7D, F)**. Second, activating mutations in AKT signaling are associated with cortical malformations and epilepsy (*85, 86*), which partially overlap with clinical features in patient with CELF2 mislocalization. In contrast, AKT inhibition suppresses neuronal hyperexcitability (*87*). This aligns with our observation that AKT inhibition rescues hyperactivity defects in patient-derived neurons (**Figure 7H)**. Moreover, CELF2 is a hyperphosphorylated protein with multiple predicted AKT sites (*88*). Notably, Ser28 in CELF1, corresponding to Ser56 in CELF2, is a confirmed AKT target (*89*). Phosphorylation at this site drives CELF1 cytoplasmic translocation, while mutation that prevents phosphorylation results in nuclear localization (*89*), which is consistent with our findings. Phosphorylation has been shown to regulate CELF family members in diverse ways, depending on the sites and upstream kinases involved. For example, in type 1 myotonic dystrophy (DM1), PKC-mediated hyperphosphorylation stabilizes CELF1 proteins (*90*), while CDK4/6-mediated phosphorylation of Ser302 in CELF1 modulates its interaction with EIF2A and mRNA targets and its subcellular localization (*89, 91*). In DM1 patient-derived cortical organoids, hyperphosphorylation of CELF2 disrupts its interaction mRNAs encoding synaptic regulators, causing hyperexcitability of glutamatergic neurons (*9*). Despite the increased activity, CELF2 does not show a shift in localization within these neurons (*9*), raising the possibility that phosphorylation may promote its cytoplasmic retention, potentially contributing to the hyperexcitability phenotype. Nonetheless, it is also plausible that AKT does not directly phosphorylate CELF2 but instead acts via indirect pathways. Future studies to identify specific phosphorylation sites on CELF2 and/or other indirect pathways will help reveal these mechanisms governing CELF2 shuttling and its impact on neuronal activity.

## MATERIALS AND METHODS

### Ethics Statement

Verbal or written consent for publication has been obtained from all study subjects or their parents in accordance with the various local institutional requirements. The collection and processing of human fetal tissues are approved by Chongqing Medical University and conducted according to the University guidelines.

### Patients

Individuals with CELF2 variants were characterized after trio exome sequencing and evaluation by clinical geneticists. Trio-exome sequencing revealed different CELF2 variants along the coding and non-coding sequence of the gene associated with heterogeneous phenotypes. Family history and clinical features of all patients were obtained by the physician at the corresponding sites. Characteristics of the patients are summarized in **Table S1**.

### Animals

All animal use was approved by the Animal Care Committee at the University of Calgary and conducted under the guidelines set forth by the Canadian Council of Animal Care. All the animals were housed in groups of one to five per cage in a room maintained at 24°C with a standard 12-hour light-dark cycle and free access to water and low-fat diet food. CD1 and C57BL6 mice were used for immunohistochemical staining of Celf2 and cell culture. The morning of plug observation was designated as embryonic day (E) 0.5. *Celf2* knock-in (KI) mice harboring one *Celf2* allele with the p.R493H variant (patient variant) were generated using the CRISPR/Cas9 approach at the University of Calgary Transgenic Facility and validated by Sanger sequencing. P4, P8, and P30 Celf2 KI mice were weighed to monitor their weight gain during early postnatal development. Celf2 knock-out (KO) mice were generated by deleting exon 2 in the *Celf2* gene, flanked by loxP sites, by crossing a CMV-Cre deleter with a conditional KO line (*Celf2^+/fl^*) (EMMA: 11977). Transgenic mice were backcrossed for five generations before use. For inhibitory neuron visualization in the brain of developing mice, Slc32a1-IRES-Cre driver mice (JAX: 016962) and Ai14(JAX: 007914) reporter mice were crossed to obtain double heterozygous mice for further analyses.

### Patient-derived iPSCs and isogenic controls

The University of Calgary (REB22-1105) approved all patient-derived cell use. Patient-derived iPSCs and isogenic controls were grown on Matrigel®-coated (Corning, USA) plates. Cells were cultured in mTeSRTM1 or mTeSR Plus^TM^ (STEMCELL Technologies, Vancouver). StemPro^TM^ Accutase^TM^ Cell Dissociation Reagent (ThermoFisher, USA) and 1:1000 ROCK inhibitor (STEMCELL Technologies, Vancouver) in mTeSRTM1 or mTeSR Plus^TM^ (STEMCELL Technologies, Vancouver) were used during cell passaging. All cells were cultured in a humidified incubator at 37°C, 5% CO2. To generate NPCs, iPSCs were grown and differentiated using the STEMdiff SMADi Neural Induction Kit (STEMCELL Technologies, #08591), following the manufacturer’s instructions. Briefly, iPSCs were seeded onto Matrigel-coated 35 mm dishes (Corning) at an initial density of 2 million cells per dish, with cell density optimized for each cell line. Cultures were rinsed with DMEM-F12 supplemented with 15 mM HEPES prior to media changes. Cells were cultured under sparse conditions to facilitate differentiation, with passaging performed at a ratio of 1:3 to 1:6. NPCs were maintained in neural progenitor media KnockOut DMEM-F12 (ThermoFisher, 12660012) supplemented with N2 Supplement (Gibco, 17502-048), B27 Supplement (ThermoFisher, 17404044), EGF (Sigma, E9644), FGF2 (Corning, 354060), and Laminin (VWR, CACB354232) beginning at passage three. For long-term storage, NPCs were dissociated into single cells using Accutase and neutralized with DMEM-F12 supplemented with 15 mM HEPES. Cell suspensions were centrifuged at 300 × g for 5 minutes, and the pellets were resuspended in cold STEMdiff Neural Progenitor Freezing Media (STEMCELL Technologies, #05838) at a concentration of 2– 3 million cells per mL and kept in liquid nitrogen. To generated neurons from NPCs, cryopreserved NPCs were thawed by rapid immersion in a 37°C water bath, pelleted, coated on Matrigel-coated 35 mm dishes, and differentiated to neurons using neuronal maturation media (BrainPhys Medium (STEMCELL Technologies, 08605) supplemented with N2 Supplement, B27 Supplement, BDNF (Genescript, Z03208), GDNF (Genescript, Z03387), and Laminin). Upon reaching 70–90% confluency, media was replaced by Neuronal Maturation Media. Half-media changes were performed every 2–3 days. Neuronal processes, including dendrites and axons, became visible within two days of media transition.

### HEK293 cells and mouse neuronal culture

HEK293 cells were cultured using Dulbecco’s Modified Eagle Medium (DMEM) (Gibco, 11995065) with 10% FBS (Multicell, 098150) and 1% penicillin/streptomycin (P/S). Each well was transfected with 1 μg of plasmid DNA (Invitrogen) using Lipofectamine™ 3000 (Thermo Fisher Scientific), following the manufacturer’s protocol. E16.5 embryos from CD1 mice were used for primary neuronal culture. Cortices were dismantled from meninges, dissociated in 0.05% trypsin-EDTA and cultured in the presence of neurobasal medium (Gibco, 21103049) supplemented with 2% B27 Supplement, 0.5 mM L-glutamine (Sigma, 56-85-9), GDNF, and 1% P/S. Neurons were plated on coverslips pre-coated with laminin (5 mg/mL) and poly-D-lysine (1 mg/mL) overnight. To prevent glial proliferation, 5 uM Arabinosyl Cytosine (Ara-C, Sigma, C6645) was added to the medium in the first day of culture (DIV1). Half of the media was changed every 2-3 days for optimum growth of neurons. For the tetrodotoxin (TTX, Sigma, T8024) treatment, final concentration of 1uM was applied to cultured neurons 48 hours prior to the harvest or fixing the cells (13 days in vitro: 13 DIV). HEK293 cell lines expressing either wildtype or mutant (R473H) CELF2 were engineered utilizing the pInducer20 expression plasmid (Addgene, #44012) and a 2nd generation lentiviral vector packaging system. This system consisted of the pCMV-VSV-G envelope plasmid (Addgene #14888) and the pCMV-dR8.2 dvpr packaging plasmid (Addgene, #8455), along with EGFP-CELF2 R473H or EGFP-CELF2 WT transfer plasmids containing either wild-type (WT) or mutant (R473H) CELF2 sequences. Lentiviral plasmids were transfected into LentiX packaging cells (Takara) using lipofectamine (Thermo) according to the manufacture’s protocol. After 48 hours, the media was harvested, filtered using a 0.45 μm syringe filter, and subjected to ultracentrifugation. The final purified lentivirus was resuspended in DMEM and utilized for the transduction of HEK293 cells.

### Antibodies and primers

Antibodies that were used are listed in **Table S3**, whereas primers are listed in **Table S4**.

### Molecular cloning and plasmids preparation

CELF2 variants were generated using the In-Fusion mutagenesis approach using In-Fusion® Snap Assembly kit (Takara Bio) with pEGFP-CELF2 WT as the template (*7*). Splicing variant reporters comprising an EGFP sequence followed by the relevant exon regions and the first and last ∼350bp of the corresponding introns in between exons were synthesized by GeneArt (ThermoFisher) or Vectorbuilder (Chicago, IL, USA) and cloned into a mammalian expression vector (pcDNA3) containing a CMV promoter to drive the expression of the reporter gene. For p.Q34X, exons 2 and 3 were included; for p.Q230X, exons 7 and 8/9 were included; and for G414AfsX45, exons 11/12 and 13 were included. Plasmids were transformed into chemically competent E. coli DH5α cells via heat shock and plated onto LB agar plates containing kanamycin. Resulting colonies were screened by colony PCR, and positive clones were confirmed by Sanger sequencing to verify correct insertion and orientation of the gene variants.

### DNA extraction and PCR

DNA extraction from cells and tail tissues for genotyping was performed using the E.Z.N.A. ® Tissue DNA Kit (Omega Bio-tek, Inc., USA) according to the manufacturer’s protocol. For each 10 µL PCR reaction, 0.5 µL of template DNA, 0.5 µL of each primer, and 5 µL of DreamTaq Green PCR Master Mix (2X) (Thermo ScientificTM, USA) was used. PCR amplification was performed by initial denaturation step at 95°C for 3 minutes (1 cycle), followed by 35 cycles of denaturation at 95°C for 30 seconds, annealing at 68°C for 30 seconds, and extension at 72°C for 30 seconds. Final extension step was carried out at 72°C for 1 minute (1 cycle). To visualize the DNA products, a 1.5% agarose gel electrophoresis was used.

### Analysis of splicing reporters

HEK293 cells and iPSCs were seeded in 12-well plates and cultured to approximately 50–60% confluency for transfection. Cells were incubated for 24 hours post-transfection before RNA extraction. Total RNA was extracted using the E.Z.N.A.® Total RNA Kit I (R6834-02, Omega Bio-tek) in accordance with the manufacturer’s instructions. The purified RNA was subsequently reverse transcribed into cDNA using the iScript™ cDNA Synthesis Kit (170-8891, Bio-Rad), following the manufacturer’s protocol. Quantitative real-time PCR (qPCR) was performed using the CFX96 Connect Real-Time PCR Detection System (Bio-Rad) with PerfeCTa® SYBR® Green FastMix® (95072-250, Quantabio). The amplification protocol consisted of an initial denaturation at 95 °C for 2 minutes, followed by 40 cycles of 94 °C for 15 seconds, 60 °C for 15 seconds, and 72 °C for 15 seconds. A subsequent melt curve analysis was performed with steps at 95 °C for 10 seconds, 65 °C for 5 seconds, and 95 °C for 0.5 seconds to confirm amplicon specificity. Each sample was analyzed in technical triplicates. Cycle threshold (Ct) values were averaged and used for relative quantification. GFP expression was normalized to the corresponding neomycin resistance gene (Neo) Ct values, with glyceraldehyde-3-phosphate dehydrogenase (GAPDH) included as an additional internal control. Relative expression levels were calculated as fold changes over the average Ct values.

### High-throughput drug screening

EGFP-CELF2 R473H HEK293T cells in DMEM (Gibco) supplemented with 10% FBS (Thermo) were seeded at 20,000 cells per well in black optical 96-well plates (COSTAR 3603) and incubated with 1 μg/mL Doxycycline overnight at 37°C in 5% CO_2_. Compounds from the FDA-approved Drug Library (Selleck Chemicals-L1300, 1406 small molecules) were then added to a concentration of 1 μM (1% DMSO) and cells were incubated an additional 24 hours. Finally, cells were stained with nucBlue (Thermo) according to the manufacturer’s directions, washed with PBS and fixed with 4% paraformaldehyde in PBS. All drug screens were conducted in duplicate. Imaging of the cells was carried out using an automated confocal laser microscope (InCell 6000, Cytiva) at a magnification of 40x. Wide-field images were acquired using the blue laser (588 nm) with a 3-second exposure at 100% laser power with the FITC (525/20 nm) emission filter. NucBlue images were obtained using the UV laser (405 nm) with a 1-second exposure at 100% laser power and the DAPI (455/50 nm) emission filter. Brightfield images were captured with a 0.06-second exposure. A total of 36 images were acquired per well, consisting of 12 fields with 3 Z-slices separated by 2 μm.

The acquired images were analyzed using FIJI Image J software, and Mander’s co-localization values were calculated by determining the ratio of Hoechst pixels that co-localized with EGFP (M2). A minimum of six images were analyzed per drug, per replicate. Candidate compounds (e.g., Triciribine, Omipalisib) were selected based on outcomes from high-throughput drug screening performed in HEK293 cells. Each drug was initially dissolved in dimethyl sulfoxide (DMSO; BioShop Canada Inc.) to generate 100 μM stock solutions, from which working concentrations were prepared. Prior to application, drugs were diluted in mTeSR™1 medium (STEMCELL Technologies, Vancouver). DMSO vehicle controls were prepared by diluting equivalent concentrations of DMSO in mTeSR™1. Media-only controls (mTeSR™1) were also included. All drug treatments were performed in duplicate. Drug concentrations (5µM – 200pM) were selected based on values reported in the literature and adjusted to minimize cytotoxicity.

Cytotoxic effects were evaluated using the CytoTox 96® Non-Radioactive Cytotoxicity Assay Kit (Promega), following the manufacturer’s instructions. For screening, iPSCs were cultured on Matrigel®-coated (STEMCELL Technologies, Vancouver) 24-well plates containing 12 mm glass coverslips (VWR, Canada) and grown to approximately 50–60% confluency. Immediately prior to treatment, spent media were removed and replaced with fresh media containing the appropriate drug, vehicle, or media-only control. Cells were incubated with treatments for 24 hours. Following incubation, cells were fixed, stained, and imaged within 48 hours. Subcellular localization of CELF2 was analyzed using FIJI (ImageJ) software as described above.

### Cell fractionation

To separate cytoplasmic and nuclear fractions, cells were harvested and centrifuged in PBS. For lysates, cells at this stage were resuspended in Laemmli Buffer (0.25M Tris-HCl (pH 6.8), 8% SDS, 40% glycerol, 0.05% Bromophenol Blue, and 100mM DTT). To obtain cytoplasmic fraction, cells were resuspended in 0.05% NP-40 Alternative (MilliporeSigma, USA), incubated on ice for 2-3 minutes and subjected to a brief centrifugation at 2000 g for 15 seconds. The supernatant was collected as cytoplasmic fraction. The pellet was washed in the same buffer twice more and was collected as nuclear fraction. The resulting cell fractions were transferred to separate tubes and mixed with Laemmli Buffer. For cortices, carefully dissected tissues were dissociated using trypsin-EDTA for 2-3 minutes and triturated prior to PBS wash and NP-40 addition. The total, cytoplasmic and nuclear fractions were subjected to sonication twice for 5 seconds each, and all samples were subsequently boiled for 1 minute and analyzed using Western blotting.

### Immunoblotting

Samples were mixed in Laemmli Buffer, boiled and loaded onto a 4-15% SDS-PAGE gel (BioRad, USA) and subjected to electrophoresis for a duration of 1.25 hours. Following electrophoresis, proteins were transferred onto a nitrocellulose membrane (Cytiva, USA). The membrane was then blocked with a solution containing 4% w/v skimmed milk powder or Bovine Serum Albumin (BSA) in TBS-T for 1 hour at room temperature. Membranes were probed overnight at 4°C with the specified primary antibodies in blocking solution. The following day, the membranes were incubated with HRP-conjugated secondary antibodies diluted in blocking solution for 1 hour at room temperature. To visualize protein bands, membranes were exposed to Western blotting detection reagent (RNP2106, Cytiva, USA) or SuperSignal West Pico PLUS Chemiluminescent Substrate (ThermoFisher, USA and imaged using the Amersham Imager 600 (GE HealthCare) and quantified using ImageJ Software.

### Immunocytochemistry

Cultured cells (HEK293, iPSCs, NPCs, or neurons) on 12 mm coverslips (VWR, Canada) were washed with PBS and fixed for 10 minutes in 4% paraformaldehyde (PFA) in PBS. PFA was then removed, and wells were rinsed twice with PBS followed by permeabilization using 0.3% TritonTM-X-100 (Sigma-Aldrich, USA) in PBS for 10 minutes. Wells were then rinsed with PBS and incubated in blocking buffer (10% Normal Donkey Serum (NDS) (Sigma-Aldrich, USA), 1% BSA (Sigma-Aldrich, USA) for 45-60 minutes at room temperature. Blocking buffer was then removed and coverslips were placed in a humidified staining chamber. Primary antibodies were diluted with an equal volume of blocking buffer and PBS and added to each coverslip. Coverslips were then incubated at 4°C overnight.

The following day, coverslips were rinsed thrice with PBS. The appropriate secondary antibodies diluted in PBS were added and incubated in the dark for 1 hour at room temperature. Nuclei were then stained using Hoechst 33258 (Sigma-Aldrich, USA). Following the incubation period, and three washes, coverslips were mounted on Fisherbrand^TM^ Superfrost^TM^ Plus Microscope Slides (Fisher Scientific, USA) using Aqua-Poly/Mount and sealed with clear nail polish.

### Immunohistochemistry

For immunostaining of embryonic or postnatal cortices, brains were dissected in ice-cold HBSS, fixed in 4% PFA at 4°C for 4 hours, cryopreserved with 30% sucrose and stored on embedding molds at -80°C prior to cryo-sectioning. Brains were cryo-sectioned coronally at 16 µm. For immunostaining of adult cortices, mice underwent perfusion fixation using PBS and 4% PFA, respectively, and brains were dissected and fixed in 4% PFA in the fridge overnight, followed by cryopreservation. Brains were cryo-sectioned coronally at 20 μm. Sections were washed in PBS twice and blocked with 5% BSA in PBS at RT with 0.3% Triton X-100. Sections were washed and incubated with appropriate primary antibodies in blocking buffer overnight at 4°C. Sections were washed thrice, followed by incubation with the appropriate secondary antibodies in diluted blocking buffer at room temperature for 1 hour.

Nuclei were counterstained with Hoechst 33258. For human tissues, embryos at 14 gestational weeks were obtained following elective termination of pregnancy and subjected to paraffin sections. The sections were dewaxed in xylene at 60°C for 1 hour and then rehydrated using a gradient alcohol series (100%-50%). Brain tissue sections were placed in 1× sodium citrate buffer (Beyotime) and boiled at 95°C for 15 minutes for antigen retrieval. After cooling to room temperature, 3% Hydrogen peroxide was added to the brain sections to block endogenous peroxidase activity. Following PBS washing, the sections were blocked with blocking buffer containing 5% normal goat serum and 0.3% Triton X-100 at 37°C for 30 minutes. Then, the sections were incubated with anti-CELF2 antibody (1:200, Abcam) at 37°C for 1-2 hours. Staining was performed according to the manufacturer’s instructions for the rabbit IgG immunohistochemistry kit (Boster Biological Technology).

### RNA immunoprecipitation (RIP)

To perform RIP-seq, cortices were dissected from P0 heterozygous Celf2 KI mice, dissociated and underwent RIP using the EZ-Magna RIP-RNA Immunoprecipitation Kit according to the manufacturer’s instructions with modifications. Briefly, cytoplasmic fraction was obtained as described above and was supplemented with the protease and RNase inhibitors. A portion of this solution was used for RNA extraction and served as “input”. The rest of the supernatant was precleared with protein A/G beads and incubated with 10 µg of rabbit anti-Celf2 or normal rabbit IgG for 3 hours at 4°C. The beads were washed and RNAs were precipitated for RNA sequencing.

### RNA-sequencing and processing

Total RNA samples from three biological replicates of Input, IgG control and CELF2 immunoprecipitates from the cortex of P0 Celf2 KI mice was isolated, extracted with phenol/chloroform, and subjected to rRNA depletion using the NEBNext rRNA Depletion Kit (New England Biolabs). RNA immunoprecipitates were then converted into cDNA libraries using the NEBNext Ultra II Directional RNA library prep kit (New England Biolabs), followed by Illumina sequencing with NEXTseq 500 (75bp reads). The sequencing quality was checked using FastQC 0.11.9 [1]. Adapters and low-quality bases were trimmed using Fastp 0.23.4 [2], as needed. To address the high abundance of ribosomal RNA (rRNA) in cellular RNA, Ribodetector 0.3.1 [3] was used to identify and remove rRNA reads from transcriptomics. Transcript quantification was performed with Kallisto 0.51.1 [4], utilizing pseudoalignment against a Mus musculus reference transcriptome downloaded from the Ensembl database [5]. These read counts were used as input for differential expression analysis. The nine sample files are available through GEO.

### Differential expression and bioinformatics

Focusing on protein-coding genes, we converted transcript counts into gene-level counts and retained only genes with identified protein-coding functions for downstream analysis. Besides, genes with low expression levels were filtered out based on Counts Per Million (CPM) threshold. Trimmed mean of M values (TMM) [6] was used as the normalization method. Principal component analysis (PCA) was performed using DESeq2 (v1.46.0) [7] on regularized log-transformed gene-level counts to assess sample reproducibility and distinguish between experimental conditions. Differentially expressed genes (DEGs) were identified using generalized linear models in edgeR (4.4.0) [8], comparing CELF2 versus Input and IgG. For this analysis, P-values were corrected for multiple comparisons using the Benjamini-Hochberg method [9] to control the false-discovery rate (FDR) at 0.01. DEGs were selected based on a minimum two-fold change in expression.

A total of 1,192 genes were identified as overexpressed in the CELF2 group. Gene ontology (GO) enrichment analysis among them was conducted using g:Profiler2 (*92*) with term size 1-2000, revealing associations with Biological Process (BP), Cellular Component (CC) and Human Phenotype Ontology (HPO) (Figure S5E, 5B). Among the identified terms, seizure-related HPO categories were highly consistent with clinical presentations observed in patients. Heatmaps (Figures 5C, F, H) were generated using z-transformed log-CPM expression values of the selected functional genes. We compared DEGs with Tier 1 seizure-related genes proposed by Macnee et.al [11], identifying 142 overlapping genes. The empirical cumulative distribution function (eCDF) of these overlapping genes, along with all DEGs, is presented in Figure 5SC. The boxplot in Figure 5D illustrates the log-fold change (LFC) distribution of these overlapping genes across various categories. Groups with significantly enriched genes, identified using the Mann–Whitney U test [12], are highlighted in yellow. The 514 overlapping DEGs genes (Tier 2) [11] were classified into five groups based on LFC of CELF2 versus IgG. Gene Ontology (GO) analysis on each subgroup was performed using DAVID [13], [14] (Figures 5E and S5D). The identified DEGs were compared with dendritic mRNAs from hippocampal CA1 neurons, as identified in [15]. The 185 overlapping genes were analyzed using ShinyGO [16], [17], [18]. (Figures 5G and S5H). DEGs were also compared with the reported dendritic mRNAs [19], revealing 115 overlapping genes (Figure S5F). GO term enrichment analysis of these shared genes, performed using the g:Profiler database [10] with a term size range of 1–2000 (Figure S5G).

### Microscopy and quantification

For microscopy and quantification, Z-stacked images of immunostained cells or sections were acquired using an Olympus FV3000 confocal microscope with a 20X or 40X or 60X objective. The images were then projected on maximum intensity using ImageJ Software (National Institutes of Health (NIH)). All images were processed using Olympus FluoView^TM^ Software and ImageJ Software. To quantify the distribution of CELF2 protein, the “Straight Line” function in ImageJ was utilized to draw a line extending from the center of the cell to the borders of the cytoplasm. A plot profile was generated for each channel. The ratio of CELF2 protein distribution in the cytoplasm versus the nucleus was calculated using Hoechst staining, which served as a marker to define the boundaries of the nucleus. For cell cultures, representative images were selected from at least three independent experiments that were used for quantifications. For the analysis and comparison of brain sections between transgenic and control animal groups, three to four anatomically matched sections per brain from at least three embryos or postnatal or adult for each group were imaged with a 20X/60X objective on the confocal microscope. For the c-FOS counting and Celf2 intensity measurement, the matched section of each image was divided into equal number of fixed size bins and then the intended quantification was performed. For the analysis of Celf2 localization in a compartment of a cell, z–stacked images were collected at 60X magnification on the FV3000 confocal microscope and projected with maximum intensity using ImageJ Software. All images were processed with Fluoview software (Olympus), ImageJ, and Adobe Photoshop. Pax6, Tbr2, and Hoechst staining were used to define the VZ, SVZ, and CP, respectively.

### Optogenetic stimulation

Under isoflurane anesthesia, mice were injected with an AAV viral construct encoding the light-activated ion channel channelrhodopsin-2 (pAAV-Syn-ChR2(H134R)-GFP) into the lateral EC (AP -3.2, ML 3.6, DV -4.6). Two weeks later, mice were connected to a patch cord and laser and received optical stimulation via a pulsed blue light at 0.4 mW 20 Hz and for 15 seconds per minute over a period of 5 minutes. Mice remained in their home cage for 90 minutes following stimulation and were then perfused with 4% paraformaldehyde under isoflurane anesthesia. Brains were cryoprotected in sucrose prior to freezing and cryosectioning for c-FOS immunohistochemistry.

### Fiber photometry

Surgery was performed under isoflurane anesthesia and mice were given Meloxicam analgesia prior to surgery and for 3 subsequent days. Using an automated robotic stereotaxic frame (Neurostar), a burr hole was drilled in the skull overlying the hippocampus. A pulled glass pipetted attached to a Nanoject III infusion system (Drummond Scientific) was used to inject 200 nL of virus into area CA1 (pGP-AAV-syn-jGCaMP8f-WPRE (AAV1); AP − 2.18; ML ± 2.0; DV 1.4). Following the virus infusion an optic fiber (NA 0.37, Core 200 um) was implanted at the injection site and fixed in place with a small amount of super glue. A headcap was then constructed out of black opaque dental acrylic to seal the skull and support the optic fiber. Mice were allowed to recover for 2 weeks. Photometry recordings were performed during contextual fear conditioning experiments. Mice were first habituated to connection of the optic patch cord for 3 days. Photometry recordings were performed with a Neurophotometrics FP3002 system controlled by Bonsai software. The contextual fear conditioning apparatus was controlled through ANYmaze tracking software (Stoelting) which also served to time lock the photometry signal to the behavioral recording through the delivery of a TTL pulse. Photometry recordings were acquired in 415 nm (isosbestic control) and 470 nm (GCaMP8f) channels in alternating frames at a frame rate of 20 FPS per channel. Both channels were calibrated to a power of 50 µW. Subsequent photometry analysis was performed using custom MATLAB scripts. Behavioral data was aligned to the photometry signal. The isosbestic channel was fit to a biexponential decay to compensate for photobleaching and the resulting signal was used to linearly scale the calcium-dependent 470 nm channel.

### Contextual fear conditioning

During the acquisition session, mice were transported to the testing room, connected to the photometry patch cable and were placed in the conditioning chamber. The training session consisted of a 2-minute habituation period prior to the delivery of 3, 0.5 mA shocks (2s duration) spaced 1 minute apart. Following the third shock the mice remained in the chamber for an additional minute. The mice were then returned to their home cage. The following day, mice were returned to the conditioning chamber for a retention test (5- minute duration) in which the mice were allowed to explore the chamber, and no foot shocks were delivered. Freezing (defined as the absence of all movement except respiration) was measured using ANYmaze behavioral tracking software. Photometry recordings were assessed during bouts of freezing and movement and examined for each specifically at the onset of the behavior (± 2 seconds) to standardize the bouts.

### Context discrimination task

Mice were transported to the testing room and handled daily for 5 days to habituate them to the experimenter and the testing. On the day of the experiment, mice were placed in Context I (a white circular open field) for 5 minutes. After returning to their home cage for 20 minutes, mice were exposed to context II (a black square open field) for 5 minutes and then returned to their home cage for 5 minutes before sacrifice. Following the behavioral task, the mice were sacrificed, and their brains were dissected, placed into OCT embedding compound and frozen in chilled isopentane at –70°C. Tissues were stored at –80°C until sectioning. Coronal sections (16 μm) were prepared using a cryostat and mounted on Superfrost Plus slides (Fisher Scientific, USA) prior to *in situ* hybridization (catFISH) experiments.

### Cellular compartment analysis of temporal activity by fluorescence *in situ* hybridization (catFISH)

RNAscope Multiplex Fluorescent Assay (Advanced Cell Diagnostics, USA) was employed to detect Arc and Homer1a mRNA following the manufacturer’s protocol. Briefly, sections were fixed in freshly prepared 4% PFA in PBS for 15 minutes at room temperature, followed by PBS washes. Probes specific for Arc (# 316911) and Homer1a (#423441-C2) were hybridized at 40°C for 2 hours in a HybEZ oven (Advanced Cell Diagnostics). Following hybridization, slides were washed and subjected to sequential amplification steps. DAPI was applied as a nuclear counterstain. Neuronal activation was analyzed by assessing the localization of Arc and Homer1a mRNA. Intranuclear signals were interpreted as transcriptional activity induced by environment A, whereas cytoplasmic signals corresponded to transcriptional activity from environment B. The number of Arc or Homer1a in the nuclear and cytoplasmic compartments of cells were quantified within each region of interest in the brain (CA1, CA3 and ACC). Image processing and quantification were performed using FIJI ImageJ.

### Statistical analyses

All data are presented as histograms with mean and standard deviation or SEM. Statistical analyses were performed with Microsoft Excel (Microsoft, WA) and GraphPad Prism. Two-tailed Student’s t-test were used to determine statistical significance between two groups while one-way ANOVA was used when there were more than two groups. P value less than 0.05 was considered significant throughout the study unless otherwise determined.

## Data Availability

All data produced in the present study are available upon reasonable request to the authors.

## List of Supplementary Materials

Materials and Methods Fig. S1 to S7

Tables S1 to S4

## Acknowledgments

We thank the patients and their families for participating in our study.

## Funding

Azrieli Foundation “RNA and the Brain” Grant G-2101-15624 (GY, JE) CIHR Project Grant OGB-192250 (GY, JE, MAM, MM)

One Child Every Child Strategic Catalyst Award (GY, JE, MAM, MM) Canadian Gene Cure Advanced Therapies for Rare Diseases (Can-GARD) (GY)

Clinical Sequencing Evidence-Generating Research Consortium, NHGRI U01HG007301 and NICHD 1R01HD112437 (DRL)

National Institute of Neurological Disorders and Stroke of the National Institutes of Health K12NS098482 (DC)

ACHRI Graduate Scholarship (MH) CIHR Postdoctoral Fellowship (MRA) CSM Postdoctoral Fellowship (MRA) ACHRI Postdoctoral Fellowship (MRA)

## Author contributions

Conceptualization: GY, MI, JE

Clinical assessment and human phenotyping: CQ, BK, SS, GV, AB, AV, ZS, RJL, FTMT, MT, CP, PL, MM, DL, RC, YI, DS, JC, MD

Investigation: MH, MZ, YR, YYO, LW, CJG, GW, BR

Formal analysis: MH, MRA, YR, YY, YYO, LW, GW, JE, GH, DJM, MI, JE, GY

Funding acquisition: MI, JE, GY Supervision: GY

Writing – original draft: MH, MRA, MM, MI, JE, GY

Writing – review & editing: All authors contributed to manuscript revision.

### Data and materials availability

All data are available in the main text or the supplementary materials.

